# Longitudinal profiling of circulating tumour DNA for tracking tumour dynamics in pancreatic cancer

**DOI:** 10.1101/2021.01.13.20248620

**Authors:** Lavanya Sivapalan, Graeme Thorn, Emanuela Gadaleta, Hemant Kocher, Helen Ross-Adams, Claude Chelala

## Abstract

The utility of circulating tumour DNA (ctDNA) for longitudinal tumour monitoring in pancreatic ductal adenocarcinoma (PDAC) has not been explored beyond mutations in the *KRAS* proto-oncogene. Here, we follow 3 patients with resectable disease and 4 patients with advanced unresectable disease, using exome sequencing of resected tissues and plasma samples (n=20) collected over a ∼2-year period from diagnosis through treatment to death or last follow-up. This includes 4 patients with ≥3 serial follow-up samples, of whom 2 are exceptionally long survivors (>5 years). Plasma from 3 chronic pancreatitis cases and 3 healthy controls were used as comparison for analysis of ctDNA mutations. We show that somatic mutation profiles in ctDNA are representative of matched tumour genomes. Furthermore, we detect and track ctDNA mutations within core PDAC driver genes, including *KRAS, NRAS, HRAS, TP53, SMAD4* and *CDKN2A*, in addition to patient-specific variants within alternative cancer drivers *(TP53, MTOR, ERBB2, EGFR, PBRM1, RNF43*). Multiple trackable (≥ 2 plasma) ctDNA alterations with potential for therapeutic actionability in PDAC are also identified. These include variants predictive of treatment response to platinum chemotherapy and/or PARP inhibition and a unique chromosome 17 kataegis locus co-localising with *ERBB2* driver variants and hypermutation signatures in one long-surviving patient. Finally, we demonstrate that exome profiling can facilitate the assessment of clonality within ctDNA mutations, for the determination of total ctDNA burden alongside temporal evolutionary relationships. These findings provide proof-of-concept for the use of whole exome sequencing of serial plasma samples to characterise ctDNA load and mutational profiles in patients with PDAC.

## Introduction

Pancreatic ductal adenocarcinoma (PDAC) is a leading cause of cancer deaths worldwide, with few effective treatment options and a dismal 5-year survival rate of ∼7% (1). This rate reflects the lack of cancer-specific symptoms during early disease and the propensity for vascular local growth and early distant metastasis, resulting in advanced inoperable disease at the time of diagnosis for >80% patients (2). Systemic chemotherapy is standard care for unresectable PDAC, despite the lack of clinically meaningful survival benefit for patients (1). The recent use of potent combination chemotherapies has delivered modest improvements in survival outcomes for a proportion of unresectable patients, although clinical applications are currently limited by toxicity (3). Even in patients who undergo surgery, early recurrences (within 6 months) occur in 28% of cases, attributed to the presence of micro-metastatic disease at the time of resection (4). To improve treatment efficacy and survival outcomes in PDAC, better stratification of patients and monitoring of tumour burden and responses to treatment is essential.

Tumour-derived genetic alterations have been identified and analysed through fragments of circulating tumour DNA (ctDNA) in peripheral blood, allowing for a minimally invasive approach to tumour sampling for monitoring strategies (5,6). ctDNA provides valuable aggregate information on multiple clonal subsets within primary tumours and metastases in individual patients, presenting significant advantages over invasive single-region tissue biopsies (7–9). However, the fractional abundance of ctDNA in patients with PDAC is known to be significantly lower compared to several other solid tumour types (10–12). These challenges to ctDNA detection have greatly limited its applications in PDAC. Most previous studies have focussed on the analysis of patients with advanced disease and a higher anticipated ctDNA burden, using droplet digital PCR (ddPCR) to detect *KRAS* variants or targeted sequencing of a small number of key hotspot mutations (6,13,14). These strategies have failed to adequately capture the extent of inter-tumoural genetic heterogeneity between PDAC tumours, resulting in significant variability between reported ctDNA detection rates and fractional abundances (15). Whilst the effects of sampling variation may not be pronounced in cancers with a high ctDNA burden or limited inter-tumoural heterogeneity, they can significantly impair the sensitivity for ctDNA detection in heterogenous disease and when the number of copies of mutant DNA in patient plasma is low (15). In such cases, ctDNA concentrations can go undetected even using platforms with high analytical sensitivities, simply due to absence of mutant molecules at a locus of interest within plasma volumes sampled at a particular timepoint (15).

In contrast, broader genomic interrogation using whole exome sequencing permits the analysis of larger numbers of mutations in plasma, with the potential to reduce sampling variation and increase the sensitivity for ctDNA detection in cancer types with high inter-tumoural genetic heterogeneity and/or a low ctDNA burden (15–19). Here, we investigate the feasibility of exome sequencing in plasma samples from 7 patients with localised and advanced PDAC. Using an optimised analytical pipeline, we identify and track ctDNA mutations from baseline (pre-treatment) throughout follow-up, in samples taken at clinically determined intervals after patients received treatment with surgery and/or chemotherapy (2-year window - until death (n=4) or last follow-up (n=3)). Our results demonstrate that exome analysis of serial plasma samples can enable longitudinal monitoring of ctDNA burden and actionable mutation profiles in response to treatment and/or disease progression in patients.

## Results

### Identification of somatic tumour mutations in plasma using exome sequencing

We retrospectively profiled, in a blinded manner, 20 blood samples from 7 patients with histologically confirmed PDAC; including 3 patients who underwent surgical resection (cases 45, 95, 28) and 4 patients who were diagnosed with surgically unresectable disease (cases 04, 13, 50, 51) and received chemotherapeutic treatment. Blood samples from 3 chronic pancreatitis (CP) and 5 healthy control (HC) cases were also studied as benign comparators **(Figure 1**, Supplementary figure 1a). Patients with PDAC had significantly more cfDNA than those with CP, whilst healthy controls had undetectable levels of cfDNA (Supplementary figure 1a). Patients with unresectable PDAC had more cfDNA than those with resectable disease (Supplementary figures 1a, b). Multiple serial blood samples collected at clinically determined intervals, separated by consecutive lines of therapy, were available from 5 patients, allowing longitudinal monitoring **(Figure 1)**. The partly unblinded clinical characteristics of the study patients are summarised in Supplementary Table 1.

**Figure 1.**
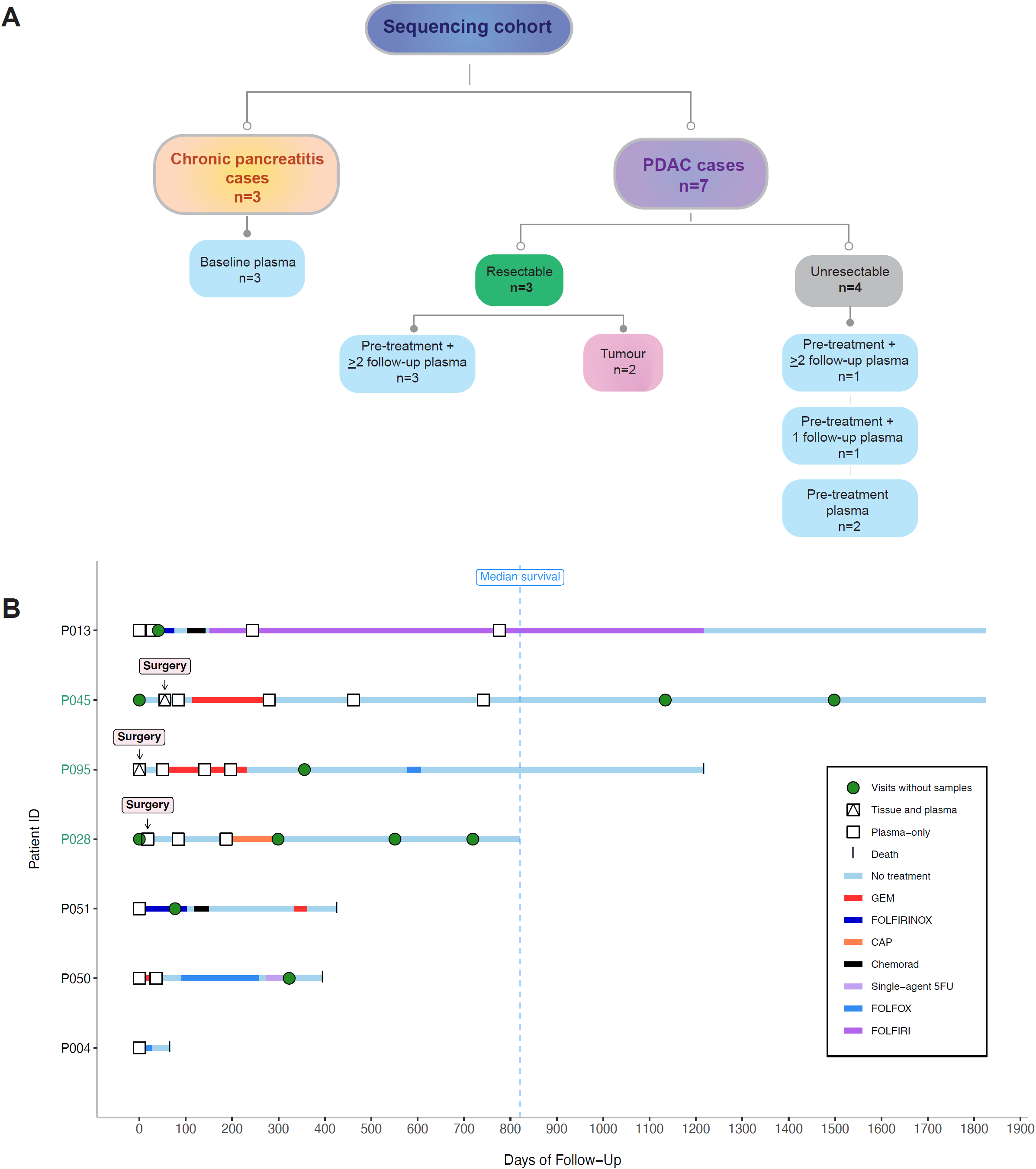
Summary of patients and samples for sequencing. **(A)** Outline of samples available for whole exome sequencing from PDAC and chronic pancreatitis (CP) control cases. **(B)** Clinical timelines including survival and treatment periods for all sequenced PDAC patients. 5FU, 5-Fluorouracil; CAP, Capecitabine; Chemorad, Chemoradiation; GEM, Gemcitabine.

Comparison between somatic mutations detected in tumour tissues and time-matched pre-treatment (P1) plasma in 2 resectable patients demonstrated a variant overlap of 43% and 31% respectively, which increased to 75% and 56% upon the comparison of tumour tissues with *combined* all time-point plasma variants **(Figure 2a, b)**. These results were used to develop and optimise our custom analysis pipeline for the identification of candidate ctDNA mutations in plasma (summarised in Supplementary figure 2; see Methods). Pathway analysis in tumour tissues and ctDNA showed enrichment of multiple well-known tumour-associated pathways (20–22), including RAS/MAPK signalling, chromatin modification, axonal guidance and DNA damage repair (DDR) across samples **(Figure 2c, d**, Supplementary Figure 3). Fragmentation profiles were inferred from plasma sequencing reads containing mutant and wild-type alleles at target loci for ctDNA. A 167bp modal fragment size was detected from most patients. Since evidence of a shortening of mutant fragment lengths (<167bp) was observed in few samples (Supplementary Figure 1c), size enrichment was not performed prior to downstream analysis of ctDNA mutations.

**Figure 2.**
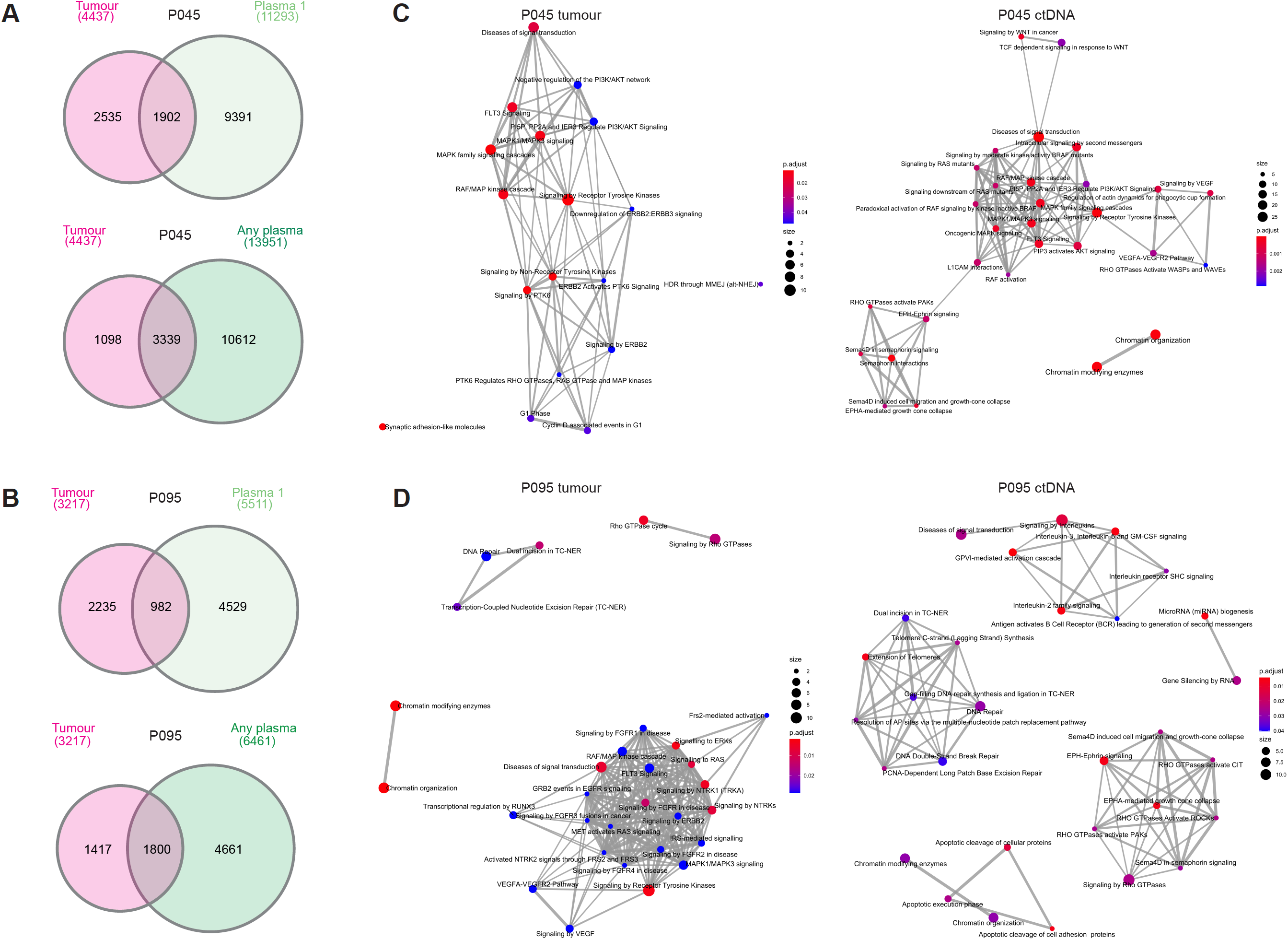
Comparison between somatic mutations in matched tumour and plasma samples from patients 45 and 95. Overlaps between somatic mutation calls in tumour tissues and baseline pre-treatment (P1) plasma *(top)*, and combined plasma (P1-P5/P4) from baseline plus follow-up sampling *(bottom)* in each patient, are shown in **(A)** and **(B)**. These comparisons were used to inform the development and optimisation of our custom analysis pipeline, for the identification of candidate ctDNA mutations in plasma. Enriched gene signalling pathways (Reactome) observed in tumour tissues and ctDNA variants from combined plasma samples are shown in **(C)** and **(D)**.

### Somatic copy number alterations are detectable in ctDNA from plasma

To determine whether somatic copy number alterations (SCNAs) observed in PDAC tumours could be detected in ctDNA, we evaluated log2 ratios and absolute somatic copy number at each mutation locus in plasma. We found similar regions of copy number (CN) gain and loss in matched tumour-plasma samples (patients 45, 95) across chromosomes 11, 15 and 18, in addition to multiple SCNAs specific to each sample type **(Figure 3a**, Supplementary figure 4). Strikingly, there were more CN calls in plasma samples compared to tumour tissues (P < 0.0001) **(Figure 3a, b)**.

**Figure 3.**
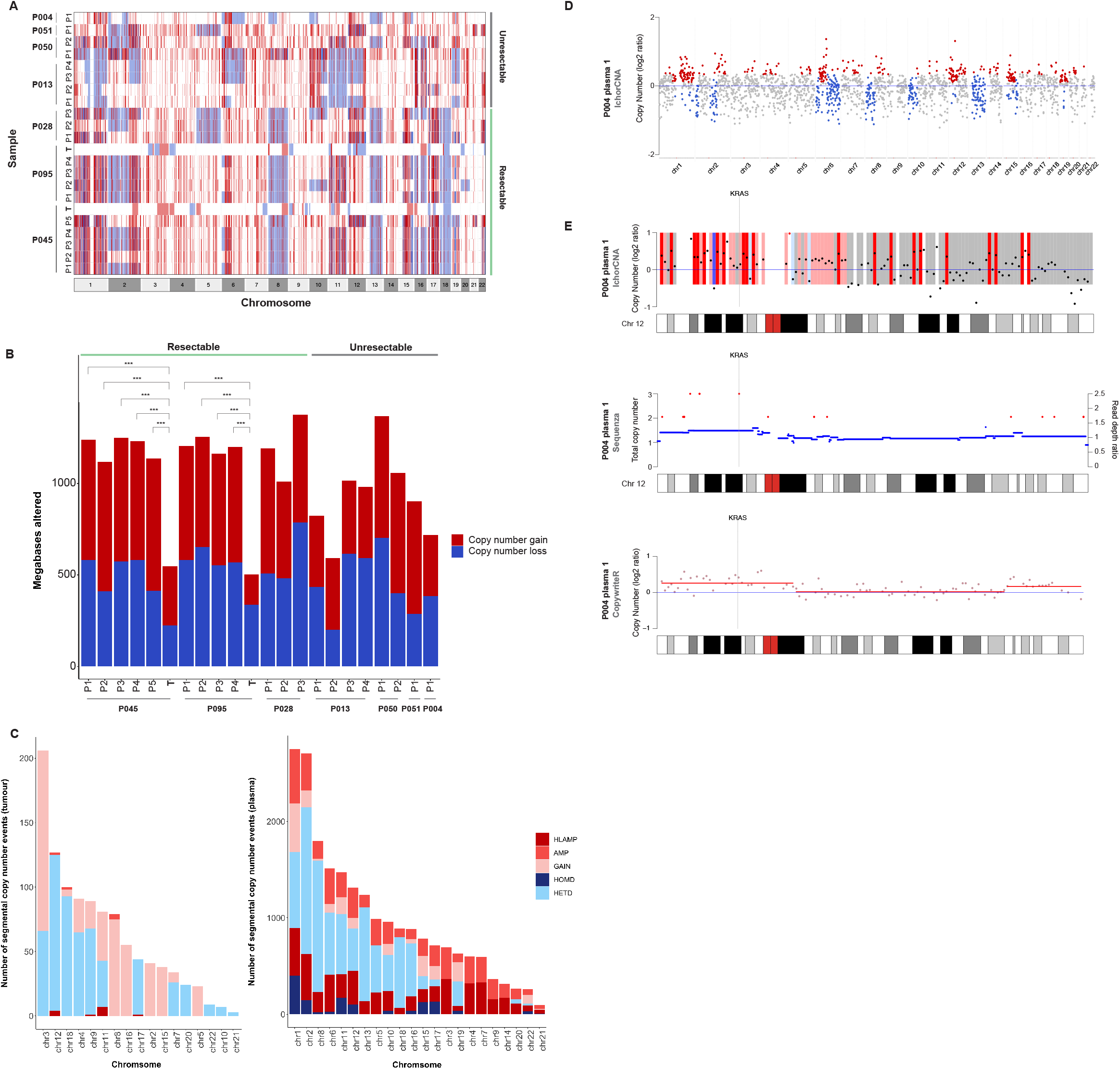
Analysis of somatic copy number alterations in tumour and plasma. Absolute copy number calls in tumour and plasma samples from the study cohort are shown in **(A)**. Gains in overall copy number are highlighted in *red* and losses of copy number are shown in *blue*. Comparison between the total number of altered copy number calls across sequenced samples is shown in **(B)**. The chi-squared test was performed for comparison (***P < 0.0001). The distribution of unique copy number events across individual chromosomes in tumour *(left)* and plasma *(right)* samples from this cohort is displayed in **(C)**, demonstrating differential enrichments for copy number gain (HLAMP, high-level amplification; AMP, amplification; GAIN, copy number gain) and loss (HOMD, homozygous deletion; HETD, heterozygous deletion) events. **(D)** The analysis of genome-wide copy number calls in plasma from one patient (patient 04) highlighted a gain *(red)* in copy number at chromosome 12p. **(E)** Further analysis of focal plasma copy number calls on chromosome 12 in this patient indicated gains in copy number at the *KRAS* locus, concurrent with the presence of *KRAS* G12D mutations. Copy number calls were determined at this region using ichorCNA *(top)*, Sequenza *(middle)* and CopywriteR (bottom).

A combined analysis of all patients identified recurrent SCNAs across several chromosomal regions, including a significant loss of copy number in both tumour (93% of all chromosome 18 tumour CN calls) and 12/20 plasma samples on chromosome 18 (82% of all chromosome 18 plasma CN calls) **(Figure 3a, c)**. We only detected CN gains in plasma from both resectable and unresectable patients on chromosomes 3, 4, 7, 9 and 14 **(Figure 3a, c)**.

We next evaluated focal SCNAs targeting key driver genes in plasma from unresectable patient 04, predicted to have the highest ctDNA burden, because of multiple liver metastases at diagnosis, alongside primary lesions in the pancreatic tail. Patient 04 displayed the poorest overall survival amongst our cohort and died <70 days after diagnosis **(Figure 1b)**. We identified distinct copy number gains on chromosome 12p, at the *KRAS* locus, in baseline plasma sampled from this patient, using three independent analysis tools **(Figure 3d, e)**. This was concurrent with the presence of detectable *KRAS* (p.G12D) mutant alleles in ctDNA. Interestingly, *KRAS* variants were not detected in plasma from any other patients.

### Plasma harbours distinct base substitution profiles and localised hypermutation

Structural variations across tumour genomes have been linked to the distribution of localised somatic hypermutation events *(kataegis)*, known to be enriched in genic regions and active chromatin elements (23,24). In light of the CN changes observed, we investigated base substitution profiles **(Figure 4a)** and the possibility for regional clustering of somatic mutations in both tumour and plasma **(Figures 4b-d)**, and found that plasma mutational profiles harboured several characteristic features. Firstly, genome-wide somatic mutational burdens in plasma were greater than in tumour tissues, consistent with the increase in arm-length SCNAs (Supplementary figure 5, **Figure 4b**). The greatest mutational burden was observed in plasma samples from patient 50 – a male in his 70’s who presented with unresectable disease at diagnosis and, unfortunately, died shortly after receiving second-line treatment (Supplementary figure 5, **Figure 4b**). Secondly, the pattern of somatic base substitutions detected in plasma exhibited a distinctive sequence context, with all samples displaying a pronounced genome-wide enrichment for C>T substitutions at Tp<u>C</u>pX trinucleotides, that was not detected in tumour tissues **(Figure 4a)**. In fact, this enrichment for C>T substitutions was also observed within multiple regions of kataegis, across plasma samples from most patients **(Figure 4b)**. We identified a unique region of kataegis on chromosome 17 in patient 45 (female in her 50’s with resectable disease), that showed a pronounced increase in T>G somatic substitution mutations in both tumour and serial plasma samples (P1-P5) **(Figures 4c, d)**. This region contained driver mutations in the *ERBB2* gene, which were detected in both tumour and in ctDNA variants from plasma, and was co-localised with larger SCNAs on chromosome 17 **(Figures 4c, d)**. This distinct chromosome 17 substitution pattern was not seen in mutation clusters from other patients (**Figure 4b)**, or in TCGA and ICGC PDAC tumour cohorts (https://dcc.icgc.org/releases/release_28/Projects/, Supplementary figure 6).

**Figure 4.**
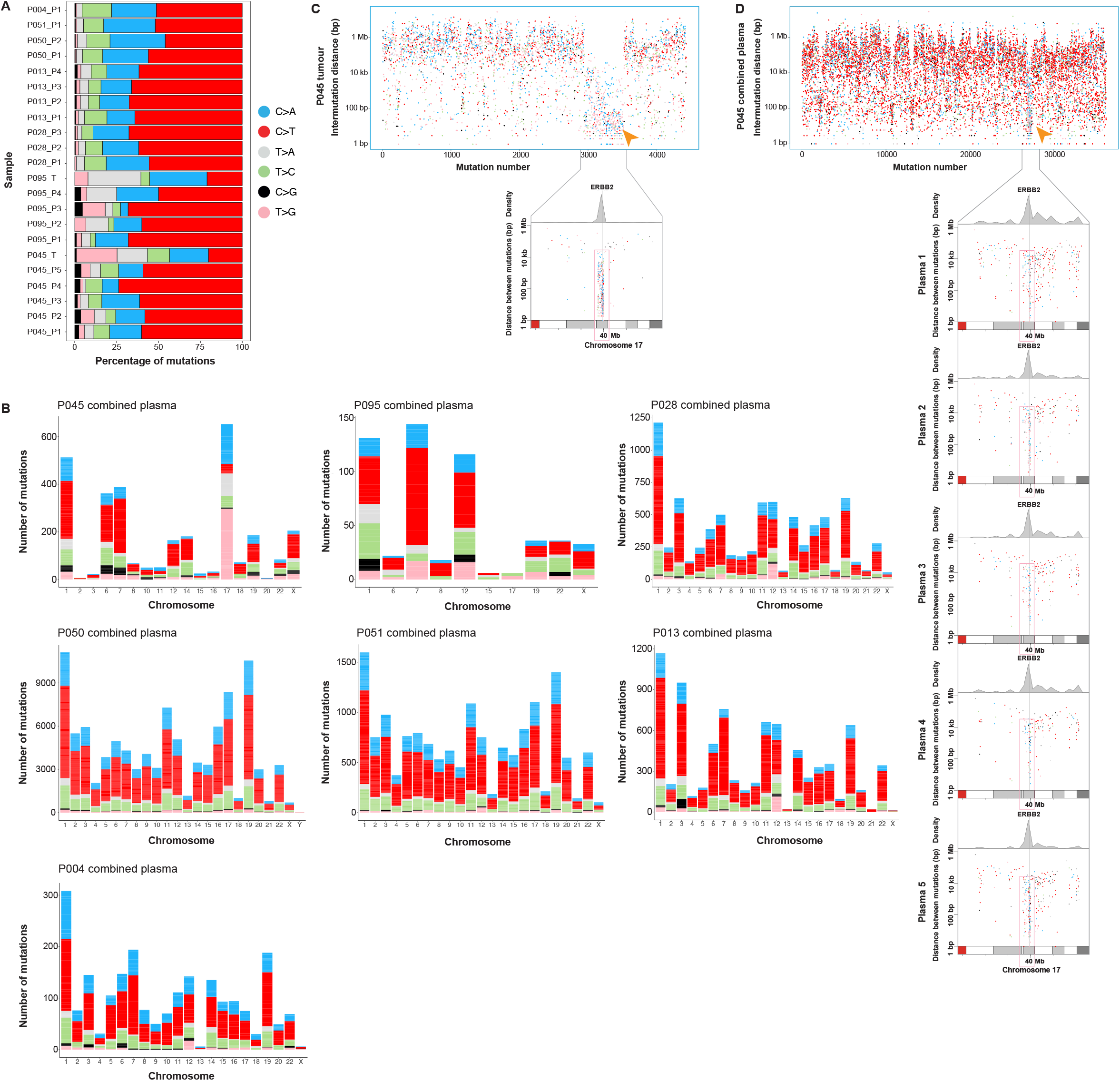
Somatic base substitution profiles and kataegis events in tumour and plasma. **(A)** Analysis of exome-wide somatic single base substitution mutations across tumour and plasma. The percentage of mutations within each sample that were characterised according to each of the 6 base substitution categories (C>T *(red)*, C>A *(blue)*, T>C *(green)*, T>A *(grey)*, T>G *(pink)*, C>G *(black)*, is displayed. A pronounced enrichment for C>T substations across plasma, compared to sequenced tumour tissues, was observed. **(B)** Bar plots showing the number of mutations within each base substitution category that were identified in regions of kataegis across individual chromosomes, in each patient. A distinct enrichment for T>G substitutions was observed within kataegis events on chromosome 17 in patient 45. **(C, D)** Rainfall plots showing the distribution of single somatic substitutions in tumour (C) and combined plasma samples (D) from patient 45, with *arrows* highlighting the presence of this unique kataegis region on chromosome 17. This kataegis region was found to contain driver mutations within the *ERBB2* gene in tumour, which were also detected in ctDNA. Inter-mutation distance is presented on the vertical axis and the number of mutations in each sample is shown on the horizontal axis.

### Clinically actionable ctDNA variants in plasma are trackable over the course of treatment in patients

The analysis of mutated genes across ctDNA highlighted multiple patient-specific variants with potential for clinical actionability. Among the variants identified in ctDNA from this cohort were missense and nonsense mutations within known PDAC driver genes, including ***KRAS*** (p.G12D), ***NRAS*** (p.T74I, p.D154Y), ***HRAS*** (p.G13C), ***TP53*** (p.E294, p.R181C, p.R196L, p.C135Y), ***SMAD4*** (p.A463T, p.R531Q) and ***CDKN2A*** (p.L130Q, p.R144H) **(Figure 5a)**. We also detected multiple patient-specific ctDNA mutations within alternative cancer driver genes, including *TP63, MTOR, ERBB2, EGFR, PBRM1, KMT2D* and *RNF43* **(Figures 5b-f)**. Most variants were detected in ≥2 serial plasma samples, with trends in variant allele fractions that were correlated with treatment and/or CA19-9 measurements **(Figures 5b-f)**. For example, in patient 13, changes in ctDNA dynamics preceded changes in CA19-9 levels in plasma **(Figures 5c, e)**.

**Figure 5.**
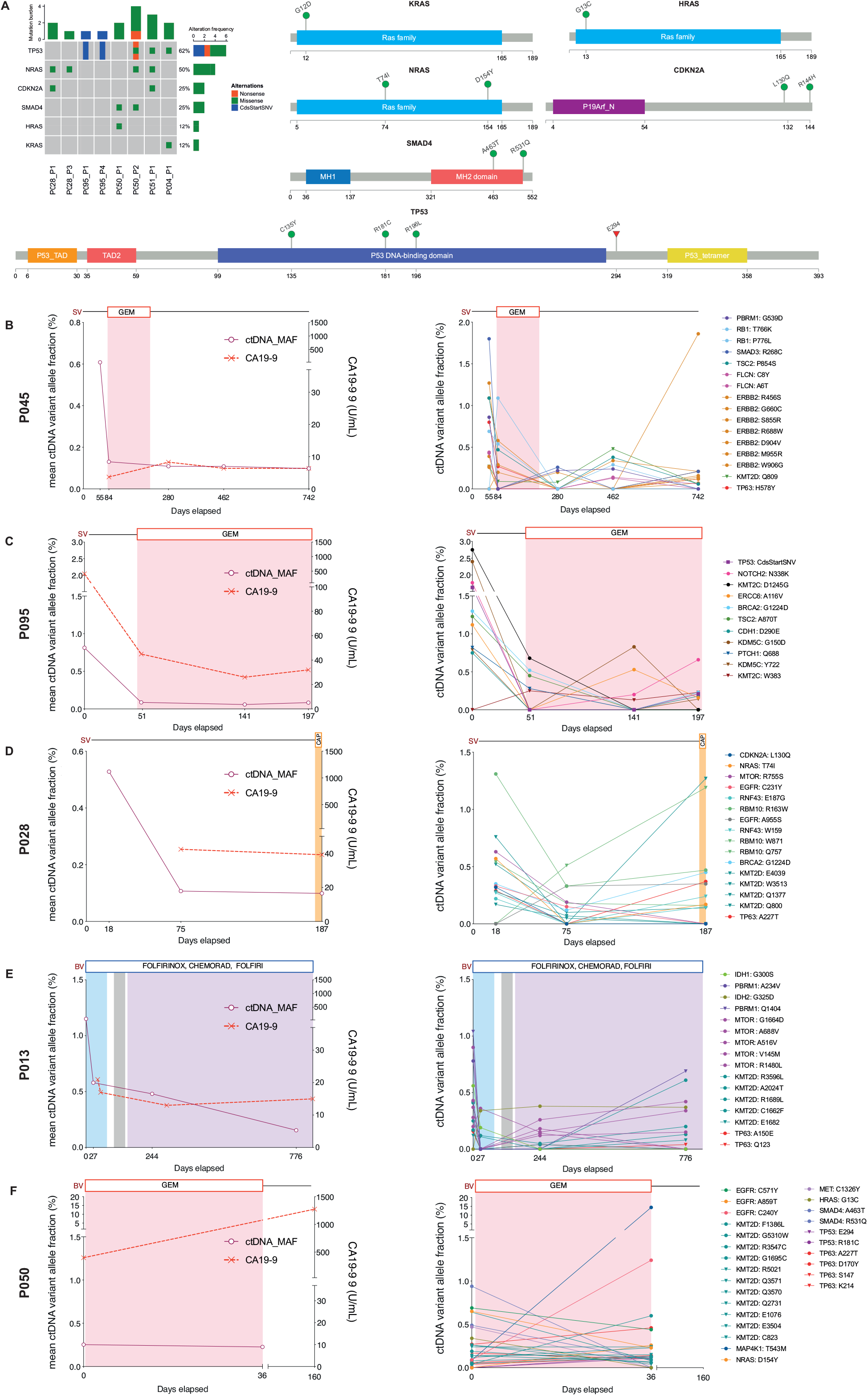
Identification of longitudinally trackable driver mutations in ctDNA. **(A)** Oncoprint showing patients with ctDNA mutations in established PDAC driver genes *(KRAS, TP53, SMAD4, CDKN2A)* and other RAS family genes *(NRAS, HRAS)* in plasma. The percentage of altered cases is displayed to the *right*. Lollipop plots displaying the mutations detected in ctDNA within individual genes are shown alongside the oncoprint. **(B-F)** In patients with multiple plasma samples, the mean mutant allele fraction (MAF) was calculated for all mutation loci in ctDNA (patient-specific plus ctDNA variants in known PDAC drivers), at each timepoint. Available measurements of CA19-9 across serial timepoints for each patient are also shown. Examples of patient-specific ctDNA mutations observed in each case are shown on the *right* (missense variants *(circles)*, nonsense variants *(triangles)*, CdsStartCNV variants *(squares)*). In two patients, temporal heterogeneity between ctDNA mutations in *RAS* and *IDH* family genes was detected **(E, F)**. CdsStartCNV; single nucleotide variant at coding start; CAP, Capecitabine; CHEMORAD (CAP), Chemoradiation (with Capecitabine); GEM, Gemcitabine.

In two patients (patients 13 and 50), we detected temporal heterogeneity between ctDNA mutations in *HRAS* (p.G13C) and *IDH*1 (p.G300S), with baseline ctDNA variants declining to undetectable levels following initial treatment in each case **(Figures 5e, f)**. However, new missense mutations in *NRAS* (p.D154Y) and *IDH2* (p.G325D) emerged in subsequent post-treatment follow-up plasma in each patient **(Figures 5e, f)**.

Using *in silico* predictions, we also identified 334 mutations in ctDNA that had been previously identified as candidates for therapeutic targeting, including 89 DNA damage-associated variants for which polyadenosine-diphosphate-ribose polymerase (PARP) inhibitor therapy or platinum chemotherapy treatment was indicated **(Figure 6**, Supplementary figure 7a). We detected a further 490 ctDNA mutations within gene signalling pathways associated with defective DNA damage repair (DDR), which amounted to a total of 187 DDR mutations that were longitudinally trackable (≥2 serial plasma samples) throughout follow-up timepoints in patients **(Figure 6)**. This included mutations in *BRCA1* and *BRCA2* genes in five patients (patients 04, 45, 50, 51, 95), as well as mutations in *PALB2* in a further three patients (patients 04, 50, 51) **(Figure 6)**. To determine whether mutations within individual DDR genes were associated with mutational signatures consistent with established modalities of genomic instability in PDAC tumours, we evaluated signature enrichments across sequenced samples **(Figure 6**, Supplementary figure 7b). In total, nine signature classes were identified in this cohort, including three that are known to be associated with mechanisms of genomic instability in tumours: double strand break repair (DSBR) *(COSMIC signature 3)*, defective mismatch repair (MMR) *(COSMIC signatures 6, 15, 20, 21, 26)* and hypermutation associated with polymerase η (POLN) *(COSMIC signature 9)* (25). **(Figure 6**, Supplementary figure 7b). Patients 45 and 95 both displayed significant enrichments for the BRCA-associated DSBR signature, which was detected in both tumour and plasma from these cases **(Figure 6**, Supplementary figure 7b). Patient 45 also displayed an abundance of tumour mutations characteristic of POLN, although to a lesser extent than the DSBR signature **(Figure 6**, Supplementary figure 7b). All patients showed enrichment for mutational signatures indicative of defective MMR in plasma **(Figure 6**, Supplementary figure 7b).

**Figure 6.**
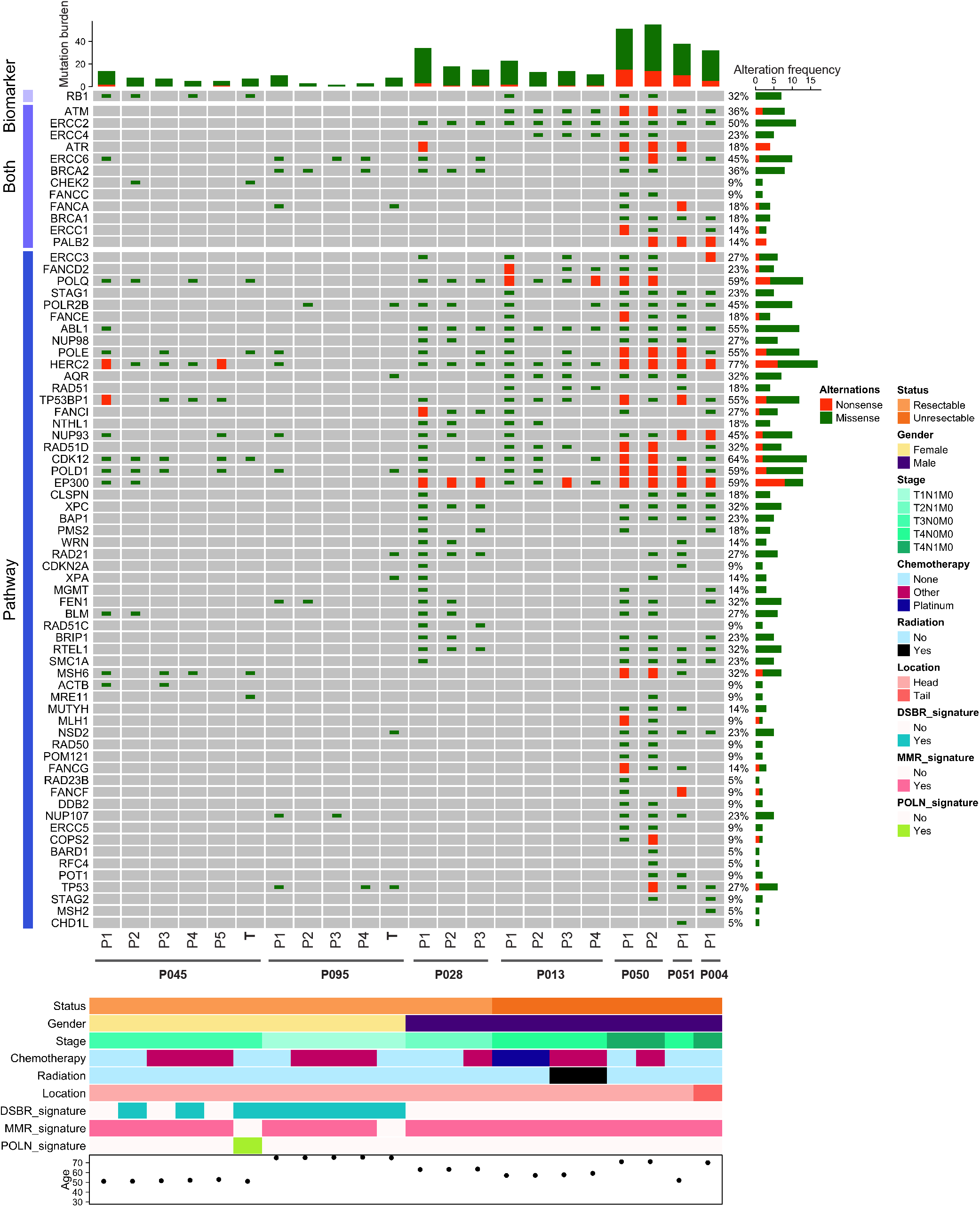
Identification of actionable therapeutic mutations in ctDNA. ctDNA mutations within genes associated with DNA damage repair (DDR) were identified across all patients from this study cohort. Oncoprint showing mutated DDR genes in ctDNA that were either predicted to confer response to platinum chemotherapy and/or PARP inhibition through *in silico* predictions (Cancer Genome Interpreter) *(Biomarkers)* or were identified within known DDR signalling pathways (Reactome) *(Pathways)*. The percentage of altered cases is displayed to the *right*. Clinical characteristics of the study cohort as well as observed enrichments for COSMIC mutational signatures associated with DDR, are shown on the *bottom* panels. Post-treatment plasma samples collected following platinum or other chemotherapies and/or radiation therapy, are indicated. DSBR, double strand break repair; MMR, mismatch repair; POLN, polymerase η (eta) hypermutation.

### Analysing longitudinal clonal evolutionary trajectories in ctDNA

Finally, we investigated whether the variant allele fractions of exome-wide ctDNA mutations could be used to determine clonal hierarchies in plasma and track clonal dynamics across serial timepoints in 4 patients (**Figure 7**, Supplementary figure 8). Utilising the temporal relationship between samples, we reconstructed longitudinally observed phylogenies from ctDNA exome data in each patient. All 4 patients displayed a good prognosis (>3 years survival) with indications of tumour response following surgical resection and/or chemotherapeutic treatment **(Figure 1b)**. Accordingly, clonal prevalence estimates of ctDNA decreased after treatment, consistent with CA19-9 measurements **(Figures 7, 5**, Supplementary figure 8). Evidence of longitudinal clonal evolution across sampled timepoints was also observed in all patients, indicating potential changes in clonal structures within ctDNA following treatment **(Figure 7**, Supplementary figure 8).

**Figure 7.**
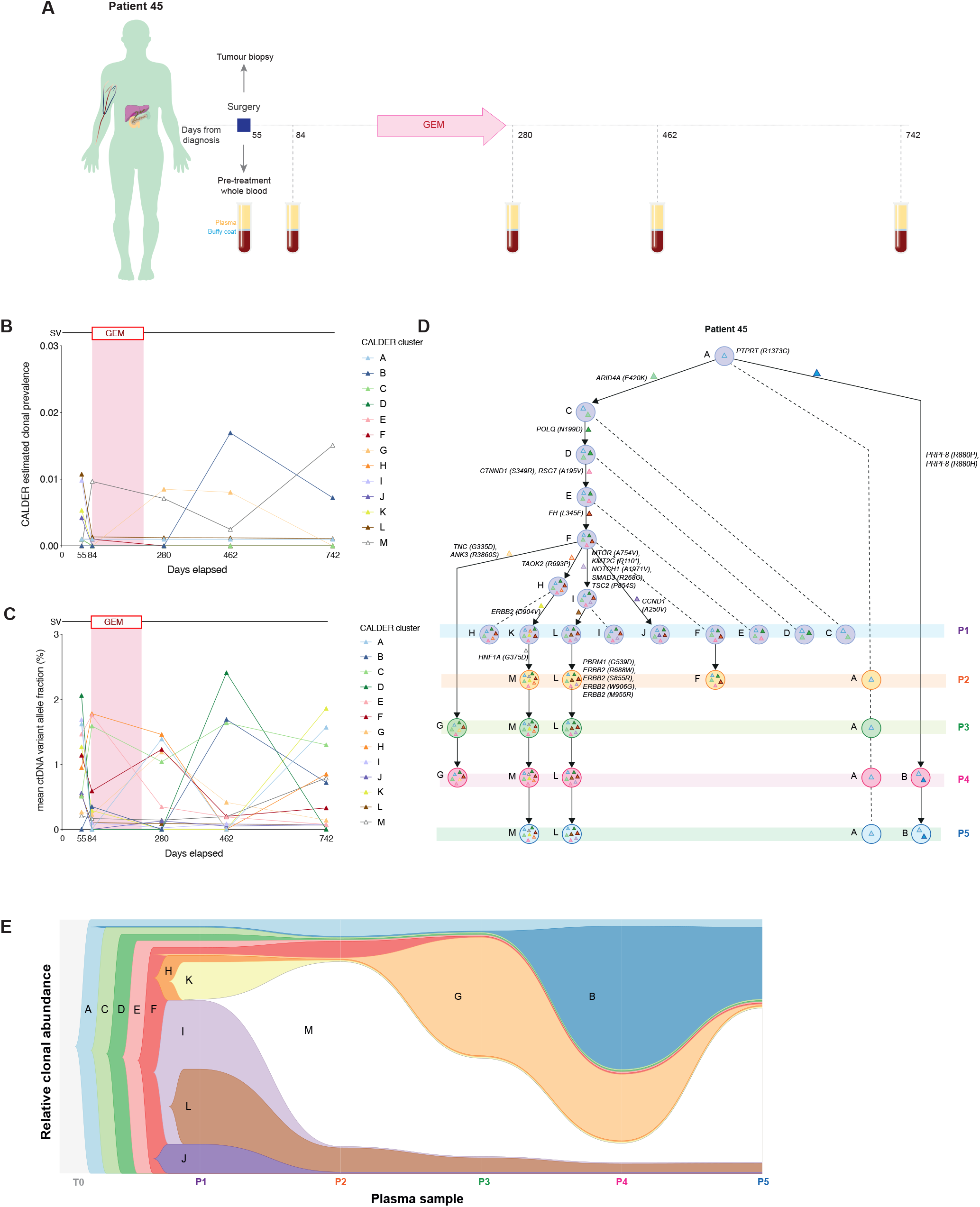
Analysis of clonal evolutionary trajectories in ctDNA from patient 45. **(A)** Clinical timeline for patient 45 showing treatment dates for surgical primary tumour resection, adjuvant chemotherapy (gemcitabine) and sampling timepoints, as days from initial diagnosis. **(B)** Scatterplot showing the total estimated prevalence of inferred clones in ctDNA, across sampled timepoints. **(C)** Scatterplot showing the mean variant allele fractions of ctDNA mutations within each inferred clone, across serial timepoints. **(D)** Longitudinally observed phylogenetic tree showing the predicted clonal evolutionary trajectories of individual ctDNA clones from patient 45. *Coloured triangles* represent mutations unique to each respective clone. Examples of unique driver mutations acquired in individual clones are shown on the tree. **(E)** Clonal diagram of the tree structure from (D) showing the differences between estimated clonal proportions across sampled timepoints in patient 45. GEM, Gemcitabine

In patient 45, who had the most extensive longitudinal follow-up amongst our cohort, with 5 serial plasma samples (>5 years survival), we detected a reduction in the number, and estimated prevalence of most ctDNA clones between pre-surgery (P1) and post-surgery (P2) samples **(Figure 7)**. Similarly, tracking clonal fractions from P3 to P5 sampling revealed a consistently low prevalence amongst most clones throughout post-treatment follow-up samples, in accordance with clinical reports of a reduction in disease burden after surgery and adjuvant gemcitabine treatment in this patient **(Figures 7b, c, e)**. This included clone L, which contained a several patient-specific *ERBB2* driver mutations (p.R688W, p.S855R, p.W906G, p.M955R), associated with the unique region of kataegis described previously **(Figures 7d, 4d)**. We also detected the emergence of new ctDNA clones (B and G) in post-treatment follow-up plasma samples, which were characterised by acquired mutations in *PRPF8* (p.R880P, p.R880H), *TNC* (p.G335D) and *ANK3* (p.R3860S) **(Figures 7d, e)**.

## Discussion

In this proof of principle study, we have demonstrated that exome sequencing of serial PDAC plasma samples and matched germline DNA can facilitate extensive characterisation of actionable mutation profiles within ctDNA, for longitudinal disease monitoring. The combination of whole exome sequencing and rigorous *in silico* filtering of plasma variants allowed for the detection of ctDNA variants in patients with both localised and advanced PDAC. Using our optimised analysis pipeline, we identified and tracked ctDNA mutations within known driver genes, in addition to variants in core PDAC signalling pathways (20,26). These results demonstrate the combined utility of broad genomic profiling with serial sampling for the identification of patient-specific ctDNA mutations that can be easily missed during targeted analysis of only a single or handful of variants amongst limited plasma sampling volumes.

We also showed that tumour-associated SCNAs could be detected in plasma and may have value for the assessment of prognosis in patients. Comparative analyses revealed the presence of several tumour-derived SCNAs in plasma from two resectable patients, in addition to SCNAs within multiple chromosomal regions that were specific to plasma. Tumour-plasma comparisons could not be made in patient 28, as the tumour tissue sample from this patient failed quality control following sequencing. Notably, a reduced overlap was observed between CNV profiles in matched tissue-plasma samples from patients 45 and 95, compared to somatic mutations. These findings are in line with recent reports, which have shown that copy number profiling has a lower overall sensitivity for ctDNA detection in cases with a low (<5%) fractional abundance of ctDNA, particularly during early or minimal residual disease, compared to somatic mutation calling (27,28). In our study, it is likely that the accurate detection of low frequency plasma copy number variants in resectable cases may have been further limited by the elevated signal-noise ratio associated with read-depth profiling in captured sequencing data (29). Although several tools have been developed and/or applied for copy number calling in ctDNA from shallow whole genome sequencing plasma datasets, their applicability for analysis of exome captured plasma data remains sub-optimal, particularly for the analysis of samples containing a low ctDNA burden (30,31). These findings highlight a pressing need for further investigation into the development of more suitable *in silico* tools for this purpose. Of note, in line with previous reports we also demonstrated copy number gains/losses and amplifications representative of PDAC tumours (32,33), as well as the prognostic value of mutant *KRAS* detection in ctDNA (14).

In addition, we identified several important and novel trends in mutational profiles within PDAC plasma samples. Genome-wide mutational burdens in plasma were significantly greater than for tumour tissues, in line with the increase in SCNAs observed. An increase in the number of predicted driver mutations in ctDNA compared to tumour tissues was also detected. As observed in tumours originating from other organs, this is indicative of the collective influence of ctDNA fragments shed from multiple different tumour clones (irrespective of anatomical biases) that are likely to capture a larger proportion of a given tumour genome, compared to a single-region biopsy specimen (16,34). Moreover, ctDNA mutational burdens can vary significantly between individuals (12). This is particularly evident in cases with early stage resectable disease, although a trend towards higher overall levels has been shown in patients with unresectable or metastatic cancers, which we demonstrate in our small cohort of patients, emphasising prognostic applications for measuring exome-wide somatic mutational burdens in PDAC ctDNA samples alongside overall isolated cell free DNA yields (12).

Profiling base substitution patterns further highlighted a distinct pattern of somatic single nucleotide substitution variants across PDAC plasma samples sequenced in this study. A pronounced enrichment for C>T transitions at Tp<u>C</u>pX trinucleotides was observed across exome-wide mutation loci in plasma, including within several regional clusters of variants with short inter-mutational distances *(kataegis)*, but was, surprisingly, absent from primary tumour tissues. Kataegis, detected across several cancer types and defined as localised hypermutation (≥6 single base somatic mutations with an average inter-mutational distance <1000bp), is characterised by a hallmark enrichment for C>T/C>G substitutions at TpCpN trinucleotides (23,24). Kataegis events were originally thought to result from aberrant APOBEC cytidine deaminase activity *(COSMIC signatures 2 and 13)*, although other mechanisms of endogenous mutagenesis have also been recently associated with kataegis events across cancers (23,24,35). We identified several regions of kataegis across plasma samples, with C>T somatic substitution variants most prevalent. However, the pronounced genome-wide enrichment for somatic C>T substitutions in plasma was not confined to such regions of localised hypermutation, indicating that this phenomenon was not solely attributable to kataegis. To date, similar genome-wide patterns of enrichment for C>T substitutions in plasma have been reported only in metastatic melanoma patients, where it has been attributed to the formation of pyrimidine dimers that arise following ultra-violet (UV) exposure (36). This has been captured through corresponding enrichments for the UV mutational signature in samples displaying this mutational profile (36). In contrast, our samples did not display any such enrichments for UV-associated mutational signatures, which have not been widely detected across PDAC tumours to date (25).

We also identified a distinct kataegis locus on chromosome 17 that was unique to one patient (patient 45). Mutations at this position (in both tumour and plasma) had a substitution profile that was dominated by T>G mutations, consistent with previous reports of an alternative kataegis signature (24). To date, only a few studies have reported on the occurrence of this rare phenomenon, which has been observed in ∼0.9% of breast cancer cases and is characterised by T>G and T>C mutations, predominantly at *N*TT and *N*TA DNA sequences (24). This distinct pattern of base substitutions most closely resembles COSMIC mutational signature 9 (POLN), attributed to polymerase η (eta) activity (24). Across our cohort, a significant enrichment for mutational signature 9 was only observed in the tumour sample from patient 45, consistent with the detection of kataegis foci displaying an alternative mutation profile in this patient. Driver mutations within this observed region co-localised with the *ERBB2* gene, and were detected in both tumour and ctDNA. Recently, D’Antonio et al (2016) demonstrated that the presence of kataegis foci on chromosomes 17 and 22 in breast tumours was associated with important clinical features and may be a putative biomarker of good prognosis in patients (35). This included an upregulation of *ERBB2* expression in patients who harboured chromosome 17 kataegis events; these patients also had a prolonged survival (35). The presence of chromosome 17 kataegis events was further associated with a downregulation of signalling pathways synonymous with aggressive metastatic tumour behaviour in breast cancer patients, including aberrations in the *NOTCH1* signalling pathway (35), as observed in ctDNA variants from patient 45. In line with these findings, we highlight that the clinical profile of patient 45 is unique, both amongst our cohort and from the typical clinical course of disease in PDAC. Patient 45 displayed an extended long-term survival exceeding 5 years from initial diagnosis, with tumour response and a reduction in overall tumour burden observed after surgery and adjuvant chemotherapy. This was followed by clinical reports of stable disease, as recorded throughout all subsequent follow-up visits for this patient, who we continue to monitor. The presence of this distinct alternative kataegis signature on chromosome 17 is a potential candidate biomarker of extended long-term survival in this patient. Whilst extensive studies in breast cancer have shown that kataegis events typically cluster around hotspot regions on chromosomes 8, 17 and 22 (35), such hotspots for kataegis events have not yet been fully characterised in PDAC. The analysis of PDAC tumours sequenced as part of the TCGA and ICGC PDAC cohorts did not reveal evidence of similar kataegis events on chromosome 17 bearing the alternative T>G substitution pattern, suggesting this phenomenon may only be present in a small sub-population of extended long-term survivors or patient-specific in nature. Amplifications of *ERBB2* in ctDNA were also reported in the recent GOZILA trial, although these too were significantly infrequent in patients with PDAC compared to other gastrointestinal cancers (37). Given the extent of inter-tumoural genetic heterogeneity between PDAC tumours, these observations merit further investigation into the pattern of mutations within kataegis foci in PDAC tumour tissues and plasma samples, to determine whether these features harbour biological and/or clinical implications for disease biology and patient survival.

We then characterised the landscape of mutated genes amongst ctDNA variants from our cohort, to determine whether variants within clinically actionable genes could be tracked over time and with treatment in patients. We identified that whilst the vast majority of individual mutations in ctDNA were patient-specific in nature, certain cancer driver genes and therapeutically relevant genes were frequently targeted by somatic mutations. We detected ctDNA mutations within all four established PDAC drivers *(KRAS, TP53, CDKN2A, SMAD4)*, in addition to variants within multiple alternative driver genes and genes associated with core PDAC signalling pathways (38–40). Whilst these findings are consistent with the extensive inter-tumoural genetic heterogeneity previously reported in PDAC tumour tissues (32), they highlight the importance of broad genomic profiling to provide a comprehensive characterisation of patient-specific ctDNA variants.

Furthermore, our results highlighted significant longitudinal changes in the variant allele fractions of ctDNA mutations. In 4 patients with ≥3 serial plasma samples, trends between mean ctDNA fractional abundances across plasma timepoints were correlated with measurements of the tumour marker CA19-9 in blood. In 3 of these patients who underwent surgical primary tumour resections, significant reductions in mean ctDNA mutant allele fractions (MAFs) were observed following surgery. Similar trends were mirrored in the fourth unresectable patient (patient 13), where a pronounced decrease in ctDNA MAFs was observed during the course of FOLFIRINOX treatment. In all 4 patients, ctDNA MAFs remained low throughout the period of subsequent clinical follow-up, which spanned several courses of chemotherapy and/or chemoradiotherapy, consistent with trends in CA19-9 levels. Importantly, these changes in ctDNA dynamics were identified prior to alterations in CA19-9 levels in 1 patient with complete CA19-9 data available (patients 13), indicating the sensitivity of ctDNA monitoring for tracking tumour burden. Notably, measurements of CA19-9 in patient 45 were significantly below the recommended upper limit of normal (37 U/mL), suggesting this patient may harbour a non-secretor Lewis phenotype (homozygous for non-functional se alleles) (41). Despite the absence of clinically useful CA19-9 measurements in this patient, longitudinal tracking of ctDNA levels reflected trends that were consistent with changes in clinically reported disease burden. These improvements to the sensitivity for longitudinal tumour monitoring in PDAC highlight potential for ctDNA tracking to manifest important changes in the clinical management of disease in patients, by enabling prompt switches between therapeutic regimens and/or management strategies prior to recurrence or disease progression.

In two patients, we detected ctDNA mutations within *RAS* and *IDH* driver genes, which demonstrated opposing mutation dynamics across serial samples, suggestive of longitudinal subclonal diversity. Matched analysis of these variants across serially sampled tumour tissues will be required to ultimately verify whether these changes are reflective of underlying molecular heterogeneity within tumour lesions in these patients. We further note that whilst mutations in isocitrate dehydrogenase (*IDH*) genes have not been extensively characterised in PDAC tumours, recent studies have highlighted the targetable potential of tumour mutations within *IDH* genes for molecularly determined treatment in PDAC (42). This indicates a promising opportunity for targeted therapy that can be detected and monitored through ctDNA and warrants further study.

Importantly, we also detected multiple ctDNA variants within genes that were either associated with DDR signalling pathways or were predicted to be potential biomarkers of response to platinum chemotherapy or PARP inhibitor agents. DNA damage repair pathways play a fundamental role in preserving genomic integrity following a plethora of insults from both exogenous and endogenous sources (43). Mutations impairing the function of genes within DDR pathways have been shown to promote a defective DNA damage response in PDAC tumours, particularly in response to double-strand DNA breaks (43). This leaves mutated tumours vulnerable to the DNA damage induced by platinum chemotherapies and PARP inhibitors (in the form of intra-strand crosslinks, or single-strand breaks leading to stalled replication forks and double strand breaks) (44,45). Whilst previous studies have assessed the detectability of germline *BRCA* mutations in ctDNA from patients with advanced PDAC (37), our results showed for the first time that multiple therapeutically relevant somatic alterations within DDR genes are detectable in PDAC ctDNA and can be tracked over time, even in cases with early disease. This included the presence of ctDNA mutations within *BRCA1* and *BRCA2* genes in several patients - including patient 95, in whom we also detected a significant enrichment for the BRCA-associated DSBR mutational signature in both tumour and plasma. Notably, enrichments for the DSBR signature were also observed in patient 45, who did not harbour *BRCA* gene mutations. These findings are consistent with previous studies which have shown that enrichments for the DSBR signature are not always underpinned by *BRCA* gene mutations in patients with PDAC (20). Patient 95 only received a short course of platinum chemotherapy, following clinical detection of liver metastatic lesions, making it difficult to accurately assess tumour response to treatment in this patient. These results support the need for further larger, prospective and statistically robust efforts to profile actionable DDR mutations within ctDNA in PDAC and assess utility for the prediction of platinum therapy response in patients.

Finally, we evaluated whether the variant allele fractions of exome-wide ctDNA mutations could be used to determine clonal dynamics and longitudinally observed evolutionary trajectories across serially sampled timepoints. Given the low frequencies of ctDNA mutations in our cohort, we applied an optimised clustering tool to distinguish between true low frequency variants and absent variants across timepoints; an important consideration when imposing longitudinal constraints (46). Longitudinal evolutionary relationships between inferred clonal structures in ctDNA were then determined using CALDER, an algorithm that was specifically developed to leverage temporal information for phylogenetic reconstruction (46). Prevalence estimates of inferred ctDNA clones in patients were in line with previous reports of ctDNA abundances in patients with ‘low ctDNA’ cancer types, including PDAC (11,37). Consistent with previous studies, we also observed an increase in the estimated prevalence of ctDNA clones in patient 13 (who had surgically unresectable disease), compared to patients with localised resectable disease, suggesting utility for clonal inference and modelling in ctDNA for quantitative assessment of disease burden in patients (12).

Furthermore, longitudinal monitoring of ctDNA clonal dynamics across serial timepoints in plasma indicated changes in clonal prevalence and structures that were consistent with clinically reported tumour burden and CA19-9 levels in individual patients studied. The analysis of clonal evolutionary trajectories in patient 45, who had the most extensive longitudinal plasma follow-up in our cohort, demonstrated a reduction in both the number, and estimated prevalence of most ctDNA clusters, following surgical primary tumour resection. Evidence of longitudinal clonal evolution was further observed in this patient, with the expansion of new subclonal structures in ctDNA following adjuvant gemcitabine treatment. These results indicate that longitudinal monitoring of ctDNA mutation profiles may have value for informing on changes in the molecular landscape of resectable PDAC tumours following surgery and adjuvant chemotherapy treatment, as indicated in recent studies (47).

In conclusion, our findings demonstrate significant biological and clinical value for exome-wide profiling of longitudinal changes in ctDNA alterations within PDAC plasma samples. To the best of our knowledge, this is the most extensive characterisation of the landscape of somatic alterations in PDAC ctDNA to date. We have shown that broad genomic profiling can enable the characterisation of tumour mutations through ctDNA, leading to the identification of important molecular features with clinical implications for prognosis, monitoring and predicting treatment response in patients. Given the heterogeneous nature of this disease, we have leveraged genomic information from multiple analytical modalities (somatic mutations, substitution patterns, copy number profiles) and serial plasma samples, in order to reliably evaluate disease burden through ctDNA measurements and to track longitudinal changes ctDNA mutation profiles that coincide with treatment response and disease progression in patients. Critically, we have also demonstrated that actionable molecular alterations can be identified in ctDNA from PDAC plasma samples, highlighting potential for the incorporation of ctDNA analysis as an adjunct to tumour genotyping in clinical trial designs. These results have shown that patient-specific alterations in ctDNA can provide insight into opportunities for molecularly defined treatment and clinical management strategies for PDAC that would not have been identified solely through the analysis of known driver genes.

## Materials and Methods

### Patients and sample collection

Blood and tumours from patients with PDAC were obtained with written informed consent from all patients and processed by the Barts Pancreatic Tissue Bank (www.bartspancreastissuebank.org.uk, Research Ethics Committee reference 13/SC/0592, project references 2015/05/QM/CC/ctDNA, 2017/06/QM/CC/C/Blood&Tissue and 2018/15/QM/CC/E/Blood). We also evaluated single-timepoint plasma samples from three individuals with chronic pancreatitis (CP), as benign disease controls for analysis of ctDNA variants. Plasma samples from n=5 healthy controls were obtained for comparative analysis of total cell-free DNA yields, but were not sequenced due to low absolute DNA quantities.

### Sample processing and DNA extraction

Whole blood samples were collected in either 10mL Vacutainer K3EDTA tubes (BD) or in RUO Cell-Free DNA Collection Tubes (Roche) and processed for plasma isolation within a maximum of 2 hours of collection through 2 centrifugation steps, each performed at room temperature for 10 mins at 1,600g, as per BPTB standard operating procedure. Following plasma isolation, the remaining buffy coat layer (containing peripheral blood lymphocytes) was carefully isolated, avoiding plasma and the red cell layer wherever possible. Plasma and buffy coat samples were stored at −80°C until DNA extraction.

cfDNA was extracted from 1.5mL-3mL available plasma using the QIAamp MinElute ccfDNA kit (Qiagen, manufacturer’s instructions), and eluted into nuclease-free (ultra-clean) water for immediate analysis. DNA from fresh-frozen bulk tumour sections and buffy coat samples was extracted using the DNeasy Blood and Tissue kit (Qiagen, manufacturer’s instructions) and stored at −80^0^C.

### Sequencing of tumour and plasma DNA

Whole exome libraries were prepared from up to 10ng of plasma cfDNA using Rubicon ThruPLEX Plasma-Seq kits. Extracted cfDNA samples were concentrated at 30°C using a SpeedVac (ThermoFisher Scientific) prior to library preparation, where required. Exome capture of plasma libraries was performed using SureSelect XT2 v6.0 human all exon (Agilent) kits with the addition of i5 and i7 xGen Universal Blocking Oligos (Integrated DNA Technologies), in line with the manufacturer’s recommendations for compatibility with ThruPLEX libraries. Enriched plasma libraries were quantified using Qubit and pooled for sequencing on the NovaSeq 6000 (Illumina) to 1000X target depth. Plasma libraries from patients 45 and 95 (P1-P4 from patient 45 and P1-P4 from patient 95) were pooled and sequenced on a HiSeq 4000 (Illumina) to 500X target depth

Tumour and germline (buffy coat) DNA samples were sheared using sonication to a target fragment size of 200bp. Sequencing libraries were prepared from up to 100ng of sheared germline DNA using HSQ SureSelect XT2 Reagent kits (Agilent), according to the manufacturer’s recommendations. Germline DNA libraries were pooled for exome enrichment using SureSelect XT2 v6.0 human all exon kits (Agilent), as described above. Enriched germline libraries were sequenced on a NovaSeq 6000 (Illumina) to 100X target depth. Sequencing of plasma and germline DNA libraries was performed at the CRUK Cambridge Institute (Genomics Core).

Whole genome sequencing was performed on extracted tumour DNA samples from patients 45 and 95. Library preparation of up to 1µg sheared tumour DNA (using TruSeq nano DNA sample preparation kits (Illumina)), sequencing and alignment/variant calling was performed by Edinburgh Genomics. The tumour tissue sample from patient 28 failed sequencing quality control and was therefore not evaluated in this study.

### Bioinformatic analysis of sequencing data

Paired-end exome sequencing reads were aligned to the hg38 human reference genome using BWA-MEM (v0.7.15). Duplicate reads were marked using Picard (from Genome Analysis Tool Kit v4.1.3.0) and removed prior to variant calling for germline DNA samples. Duplicate reads were left unmarked for the analysis of plasma samples. Base quality score recalibration and indel realignment was performed on aligned sequencing reads using GATK v4.1.3.0.

Variants were then called per patient, using samtools (v1.9) mpileup, and VarScan (v2.4.3) in multi-sample mode, with a minimum coverage of 3 reads with one read on each strand for a variant to be called, and annotated using ANNOVAR. Called variants were subsequently filtered to remove any mutations that were absent in the COSMIC91 database but with a corresponding identifier in the dbSNP database. Variant filters were also implemented based on annotated exonic function to remove mutations with ‘synonymous’ or ‘unknown’ classifications. Only variants with an alternate allele base quality score ≥25, and no alternate reads in either matched germline DNA (at a site covered ≥20x) or plasma DNA from CP cases, were retained for further analysis.

### Filtering of plasma variants

To enrich for candidate ctDNA mutations within filtered plasma sequencing reads and minimise the number of false-positive variant calls, multi-allelic variants were removed and only mutations with a single alternative genotype across serial plasma samples from each patient were retained. Alternate allele frequencies for plasma variants were assessed across normal populations from the 1000 Genomes Project (1000G), the Genome Aggregation Database (gnomAD) and Haplotype Map (HapMap) project using our variant analysis tool (SNPnexus, http://www.snp-nexus.org/) (48). Plasma variants with reported mutant allele frequencies (>0%) across these population databases were flagged. Known or predicted (tier 1 and tier 2) driver mutations in plasma, and actionable mutations of relevance for targeted treatment, were further annotated using the Cancer Genome Interpreter (CGI) function in SNPnexus. A summary of the complete analytical pipeline used for variant calling and analysis is shown in Supplementary figure 2.

### Estimation of copy number alterations in tumour and plasma

Genome-wide copy number alterations in tumour and plasma samples were determined using the ichorCNA tool (v0.3.2). The copy number signal for each patient was modelled according to a two-component mixture, comprised of a non-tumour proportion and the proportion of tumour cells carrying a specific mutation. BAM files from paired tumour-germline or plasma-germline samples were provided as input for analysis. WIG files with non-overlapping 1Mb bins across all chromosomes were generated from matched WGS (tumour)/WES (plasma) and normal (PBMC-derived) BAM files for each patient, using the ‘readCounter’ function from HMMCopy, according to a probabilistic Hidden Markov Model (HMM). Only variant reads with a mapping quality ≥20 were used for the generation of WIG files. Aligned reads were counted based on the overlap within each bin and centromeres filtered based on chromosomal gap coordinates (including 1Mb bin up-stream and down-stream of the gap). Read counts for each bin were normalised to correct for GC content and mappability biases, using a LOESS regression curve fitting applied to only autosomes. Pathology-informed tumour cellularity estimates were used to inform copy number predictions from ichorCNA for all tumour tissue samples. Tumour fractions were estimated in plasma samples using the intrinsic purity prediction function of ichorCNA. The global optimum for estimated tumour fraction in plasma samples was initialised according to expected normal cell contamination values, in the range of (0.2, 0.35, 0.5, 0.65, 0.8, 0.9, 0.99). Analyses of both tumour and plasma samples were run using the ‘clonal-only’ mode of ichorCNA and predictions trained and analysed using only autosomes to reduce complexity. Total copy number estimates from ichorCNA were verified using the CopyWriteR Bioconductor package (v2.0.6) and Sequenza (v3.0.0).

### Identification of enriched mutational signatures in tumour and plasma

Mutational signatures in tumour and plasma were analysed using the R package deconstructSigs (v1.8.0), alongside the Bioconductor library BS.genome.Hsapiens.UCSC.hg38.

### Analysis of pathway enrichments

Enriched gene signalling pathways were analysed using ClueGO and the R package ReactomePA (v1.16.2). A hypergeometric model was used to determine whether the number of selected genes associated with each pathway in the Reactome database was greater than expected by chance. *P* values for individual pathway enrichments were calculated based on the hypergeometric model.

### Identification of kataegis events in tumour and plasma

Rainfall plots for the visualisation of inter-mutational distances across the genome were generated using the R package KaryoploteR (v1.16.0) (49). A positive kataegis event was defined as the presence of 6 or more individual mutations with an average inter-mutational distance of ≤1000bp. Quantitative analysis of kataegis events identified in rainfall plots for each sample was performed using the R packages ClusteredMutations (v1.0.1), MAFtools (v0.9.3) and Seqkat (v0.0.8) (50). The minimum hypermutation score used to classify windows in the sliding binomial test as significant during Seqkat analysis was 5, the maximum log_10_(inter-mutational distance) allowed for SNVs to be grouped into the same kataegis event was 4 and the minimum number of mutations required within a cluster to be classified as kataegis was 6.

### Inferring clonal population structures and evolutionary trajectories in ctDNA

To ensure high confidence in parameter estimates for clonal evolutionary trajectories, only 4 patients with ≥3 serial plasma samples were analysed. Filtered lists of candidate ctDNA variants, derived using the pipeline described above, were considered for clonal evolution determination in each patient. The number of reference and alternate reads for each variant for each patient per plasma sample were clustered using Absence Aware Clustering (https://github.com/raphael-group/Absence-Aware-Clustering) to statistically infer independent mutation clusters in ctDNA based on similar variant allele fractions, whilst distinguishing between mutations that were present at very low mutant frequencies and mutations that were absent at each timepoint. The clustered mutations were then run through CALDER (46), which returned clonal determinations and clonal prevalence per clone at each plasma time point. The raw data was visualised using the timescape package in R (v1.14.0), with the clonal trees outputted by timescape redrawn.

## Data Availability

Data will be submitted and available from the EGA upon acceptance.

## Acknowledgements

We are grateful to patients who donated tissues to the Barts Pancreatic Tissue Bank (BPTB) (www.bartspancreastissuebank.org.uk). We are grateful to all BPTB staff for setting up the framework for collection and distribution of samples and clinical data.

## References

1. Siegel RL, Miller KD, Jemal A. Cancer statistics, 2020. CA Cancer J Clin. American Cancer Society; 2020;70:7–30.

2. Willenbrock F, Cox CM, Parkes EE, Wilhelm-Benartzi CS, Abraham AG, Owens R, et al. Circulating biomarkers and outcomes from a randomised phase 2 trial of gemcitabine versus capecitabine-based chemoradiotherapy for pancreatic cancer. Br J Cancer [Internet]. 2020; Available from: https://doi.org/10.1038/s41416-020-01120-z

3. Conroy T, Hammel P, Hebbar M, Ben Abdelghani M, Wei AC, Raoul J-L, et al. FOLFIRINOX or Gemcitabine as Adjuvant Therapy for Pancreatic Cancer. N Engl J Med. United States; 2018;379:2395–406.

4. Tummers WS, Groen J V, Sibinga Mulder BG, Farina-Sarasqueta A, Morreau J, Putter H, et al. Impact of resection margin status on recurrence and survival in pancreatic cancer surgery. BJS (British J Surgery) [Internet]. 2019;106:1055–65. Available from: https://doi.org/10.1002/bjs.11115

5. Sugimori M, Sugimori K, Tsuchiya H, Suzuki Y, Tsuyuki S, Kaneta Y, et al. Quantitative monitoring of circulating tumor DNA in patients with advanced pancreatic cancer undergoing chemotherapy. Cancer Sci [Internet]. 2019/12/24. John Wiley and Sons Inc., 2020;111:266–78. Available from: https://pubmed.ncbi.nlm.nih.gov/31746520

6. Watanabe F, Suzuki K, Tamaki S, Abe I, Endo Y, Takayama Y, et al. Longitudinal monitoring of KRAS-mutated circulating tumor DNA enables the prediction of prognosis and therapeutic responses in patients with pancreatic cancer. PLoS One. Public Library of Science; 2020;14:e0227366.

7. Bernard V, Kim DU, San Lucas FA, Castillo J, Allenson K, Mulu FC, et al. Circulating Nucleic Acids Are Associated With Outcomes of Patients With Pancreatic Cancer. Gastroenterology. 2019;156:108-118.e4.

8. Cristiano S, Leal A, Phallen J, Fiksel J, Adleff V, Bruhm DC, et al. Genome-wide cell-free DNA fragmentation in patients with cancer. Nature. England; 2019;570:385–9.

9. Zill OA, Banks KC, Fairclough SR, Mortimer SA, Vowles J V, Mokhtari R, et al. The Landscape of Actionable Genomic Alterations in Cell-Free Circulating Tumor DNA from 21,807 Advanced Cancer Patients. Clin Cancer Res. United States; 2018;24:3528–38.

10. Abbosh C, Birkbak NJ, Wilson GA, Jamal-Hanjani M, Constantin T, Salari R, et al. Phylogenetic ctDNA analysis depicts early-stage lung cancer evolution. Nature. Macmillan Publishers Limited, part of Springer Nature. All rights reserved., 2017;545:446.

11. Mouliere F, Chandrananda D, Piskorz AM, Moore EK, Morris J, Ahlborn LB, et al. Enhanced detection of circulating tumor DNA by fragment size analysis. Sci Transl Med. United States; 2018;10.

12. Bettegowda C, Sausen M, Leary RJ, Kinde I, Wang Y, Agrawal N, et al. Detection of circulating tumor DNA in early- and late-stage human malignancies. Sci Transl Med. United States; 2014;6:224ra24.

13. Hadano N, Murakami Y, Uemura K, Hashimoto Y, Kondo N, Nakagawa N, et al. Prognostic value of circulating tumour DNA in patients undergoing curative resection for pancreatic cancer. Br J Cancer. England; 2016;115:59–65.

14. Mohan S, Ayub M, Rothwell DG, Gulati S, Kilerci B, Hollebecque A, et al. Analysis of circulating cell-free DNA identifies KRAS copy number gain and mutation as a novel prognostic marker in Pancreatic cancer. Sci Rep. Nature Publishing Group UK; 2019;9:11610.

15. Wan JCM, Heider K, Gale D, Murphy S, Fisher E, Mouliere F, et al. ctDNA monitoring using patient-specific sequencing and integration of variant reads. Sci Transl Med [Internet]. 2020;12:eaaz8084. Available from: http://stm.sciencemag.org/content/12/548/eaaz8084.abstract

16. Chicard M, Colmet-Daage L, Clement N, Danzon A, Bohec M, Bernard V, et al. Whole-Exome Sequencing of Cell-Free DNA Reveals Temporo-spatial Heterogeneity and Identifies Treatment-Resistant Clones in Neuroblastoma. Clin Cancer Res. 2018;24:939 LP – 949.

17. Beltran H, Romanel A, Casiraghi N, Sigouros M, Benelli M, Xiang J, et al. Whole exome sequencing (WES) of circulating tumor DNA (ctDNA) in patients with neuroendocrine prostate cancer (NEPC) informs tumor heterogeneity. J Clin Oncol. American Society of Clinical Oncology; 2017;35:5011.

18. Giroux Leprieur E, Hélias-Rodzewicz Z, Takam Kamga P, Costantini A, Julie C, Corjon A, et al. Sequential ctDNA whole-exome sequencing in advanced lung adenocarcinoma with initial durable tumor response on immune checkpoint inhibitor and late progression. J Immunother Cancer [Internet]. 2020;8:e000527. Available from: http://jitc.bmj.com/content/8/1/e000527.abstract

19. Murtaza M, Dawson S-J, Tsui DWY, Gale D, Forshew T, Piskorz AM, et al. Non-invasive analysis of acquired resistance to cancer therapy by sequencing of plasma DNA. Nature. England; 2013;497:108–12.

20. Waddell N, Pajic M, Patch A-M, Chang DK, Kassahn KS, Bailey P, et al. Whole genomes redefine the mutational landscape of pancreatic cancer. Nature. England; 2015;518:495–501.

21. Jones S, Zhang X, Parsons DW, Lin JC-H, Leary RJ, Angenendt P, et al. Core signaling pathways in human pancreatic cancers revealed by global genomic analyses. Science. United States; 2008;321:1801–6.

22. Bailey P, Chang DK, Nones K, Johns AL, Patch A-M, Gingras M-C, et al. Genomic analyses identify molecular subtypes of pancreatic cancer. Nature [Internet]. Nature Publishing Group, a division of Macmillan Publishers Limited. All Rights Reserved., 2016;531:47. Available from: http://dx.doi.org/10.1038/nature16965

23. Nik-Zainal S, Alexandrov LB, Wedge DC, Van Loo P, Greenman CD, Raine K, et al. Mutational processes molding the genomes of 21 breast cancers. Cell [Internet]. 2012/05/17. Cell Press; 2012;149:979–93. Available from: https://pubmed.ncbi.nlm.nih.gov/22608084

24. Nik-Zainal S, Davies H, Staaf J, Ramakrishna M, Glodzik D, Zou X, et al. Landscape of somatic mutations in 560 breast cancer whole-genome sequences. Nature [Internet]. 2016;534:47–54. Available from: https://doi.org/10.1038/nature17676

25. Alexandrov LB, Nik-Zainal S, Wedge DC, Aparicio SAJR, Behjati S, Biankin A V, et al. Signatures of mutational processes in human cancer. Nature [Internet]. 2013;500:415–21. Available from: https://doi.org/10.1038/nature12477

26. Bailey P, Chang DK, Nones K, Johns AL, Patch A-M, Gingras M-C, et al. Genomic analyses identify molecular subtypes of pancreatic cancer. Nature. Nature Publishing Group, a division of Macmillan Publishers Limited. All Rights Reserved., 2016;531:47.

27. Adalsteinsson VA, Ha G, Freeman SS, Choudhury AD, Stover DG, Parsons HA, et al. Scalable whole-exome sequencing of cell-free DNA reveals high concordance with metastatic tumors. Nat Commun. 2017;8:1324.

28. Heitzer E, Ulz P, Belic J, Gutschi S, Quehenberger F, Fischereder K, et al. Tumor-associated copy number changes in the circulation of patients with prostate cancer identified through whole-genome sequencing. Genome Med. 2013;5:30.

29. Favero F, Joshi T, Marquard AM, Birkbak NJ, Krzystanek M, Li Q, et al. Sequenza: allele-specific copy number and mutation profiles from tumor sequencing data. Ann Oncol Off J Eur Soc Med Oncol. 2015;26:64–70.

30. Scheinin I, Sie D, Bengtsson H, van de Wiel MA, Olshen AB, van Thuijl HF, et al. DNA copy number analysis of fresh and formalin-fixed specimens by shallow whole-genome sequencing with identification and exclusion of problematic regions in the genome assembly. Genome Res. 2014;24:2022–32.

31. Van Roy N, Van Der Linden M, Menten B, Dheedene A, Vandeputte C, Van Dorpe J, et al. Shallow Whole Genome Sequencing on Circulating Cell-Free DNA Allows Reliable Noninvasive Copy-Number Profiling in Neuroblastoma Patients. Clin Cancer Res [Internet]. 2017;23:6305 LP – 6314. Available from: http://clincancerres.aacrjournals.org/content/23/20/6305.abstract

32. Integrated Genomic Characterization of Pancreatic Ductal Adenocarcinoma. Cancer Cell. United States; 2017;32:185-203.e13.

33. Harada T, Chelala C, Bhakta V, Chaplin T, Caulee K, Baril P, et al. Genome-wide DNA copy number analysis in pancreatic cancer using high-density single nucleotide polymorphism arrays. Oncogene [Internet]. 2007/10/22. 2008;27:1951–60. Available from: https://pubmed.ncbi.nlm.nih.gov/17952125

34. Manier S, Park J, Capelletti M, Bustoros M, Freeman SS, Ha G, et al. Whole-exome sequencing of cell-free DNA and circulating tumor cells in multiple myeloma. Nat Commun. England; 2018;9:1691.

35. D’Antonio M, Tamayo P, Mesirov JP, Frazer KA. Kataegis Expression Signature in Breast Cancer Is Associated with Late Onset, Better Prognosis, and Higher HER2 Levels. Cell Rep. 2016;16:672–83.

36. Cutts A, Venn O, Dilthey A, Gupta A, Vavoulis D, Dreau H, et al. Characterisation of the changing genomic landscape of metastatic melanoma using cell free DNA. npj Genomic Med [Internet]. 2017;2:25. Available from: https://doi.org/10.1038/s41525-017-0030-7

37. Nakamura Y, Taniguchi H, Ikeda M, Bando H, Kato K, Morizane C, et al. Clinical utility of circulating tumor DNA sequencing in advanced gastrointestinal cancer: SCRUM-Japan GI-SCREEN and GOZILA studies. Nat Med [Internet]. 2020;26:1859–64. Available from: https://doi.org/10.1038/s41591-020-1063-5

38. Dawkins JBN, Wang J, Maniati E, Heward JA, Koniali L, Kocher HM, et al. Reduced Expression of Histone Methyltransferases KMT2C and KMT2D Correlates with Improved Outcome in Pancreatic Ductal Adenocarcinoma. Cancer Res. 2016;76:4861–71.

39. Morran DC, Wu J, Jamieson NB, Mrowinska A, Kalna G, Karim SA, et al. Targeting mTOR dependency in pancreatic cancer. Gut [Internet]. 2014;63:1481 LP – 1489. Available from: http://gut.bmj.com/content/63/9/1481.abstract

40. Lee J, Lee J, Choi C, Kim JH. Blockade of integrin α3 attenuates human pancreatic cancer via inhibition of EGFR signalling. Sci Rep [Internet]. 2019;9:2793. Available from: https://doi.org/10.1038/s41598-019-39628-x

41. Ballehaninna UK, Chamberlain RS. The clinical utility of serum CA 19-9 in the diagnosis, prognosis and management of pancreatic adenocarcinoma: An evidence based appraisal. J Gastrointest Oncol. China; 2012;3:105–19.

42. Brody JR, Yabar CS, Zarei M, Bender J, Matrisian LM, Rahib L, et al. Identification of a novel metabolic-related mutation (IDH1) in metastatic pancreatic cancer. Cancer Biol Ther. 2018;19:249–53.

43. Knijnenburg TA, Wang L, Zimmermann MT, Chambwe N, Gao GF, Cherniack AD, et al. Genomic and Molecular Landscape of DNA Damage Repair Deficiency across The Cancer Genome Atlas. Cell Rep [Internet]. 2018;23:239-254.e6. Available from: http://www.sciencedirect.com/science/article/pii/S2211124718304376

44. Pishvaian MJ, Joseph Bender R, Matrisian LM, Rahib L, Hendifar A, Hoos WA, et al. A pilot study evaluating concordance between blood-based and patient-matched tumor molecular testing within pancreatic cancer patients participating in the Know Your Tumor (KYT) initiative. Oncotarget. 2017;8:83446–56.

45. Park W, Wong W, Yu KH, Varghese AM, Riaz N, Balachandran VP, et al. Homologous recombination deficiency (HRD): A biomarker for first-line (1L) platinum in advanced pancreatic ductal adenocarcinoma (PDAC). J Clin Oncol [Internet]. American Society of Clinical Oncology; 2019;37:4132. Available from: https://doi.org/10.1200/JCO.2019.37.15_suppl.4132

46. Myers MA, Satas G, Raphael BJ. CALDER: Inferring Phylogenetic Trees from Longitudinal Tumor Samples. Cell Syst [Internet]. 2019;8:514-522.e5. Available from: http://www.sciencedirect.com/science/article/pii/S2405471219301917

47. Sakamoto H, Attiyeh MA, Gerold JM, Makohon-Moore AP, Hayashi A, Hong J, et al. The Evolutionary Origins of Recurrent Pancreatic Cancer. Cancer Discov [Internet]. 2020;10:792 LP – 805. Available from: http://cancerdiscovery.aacrjournals.org/content/10/6/792.abstract

48. Oscanoa J, Sivapalan L, Gadaleta E, Dayem Ullah AZ, Lemoine NR, Chelala C. SNPnexus: a web server for functional annotation of human genome sequence variation (2020 update). Nucleic Acids Res [Internet]. 2020;48:W185–92. Available from: https://doi.org/10.1093/nar/gkaa420

49. Gel B, Serra E. karyoploteR: an R/Bioconductor package to plot customizable genomes displaying arbitrary data. Bioinformatics [Internet]. 2017;33:3088–90. Available from: https://doi.org/10.1093/bioinformatics/btx346

50. Yousif F, Prokopec SD, Sun RX, Fan F, Lalansingh CM, Drysdale E, et al. The Origins and Consequences of Localized and Global Somatic Hypermutation. bioRxiv [Internet]. 2018;287839. Available from: http://biorxiv.org/content/early/2018/06/29/287839.abstract

