## Supplementary Figures for "Longitudinal profiling of circulating tumour DNA for tracking tumour dynamics in pancreatic cancer"

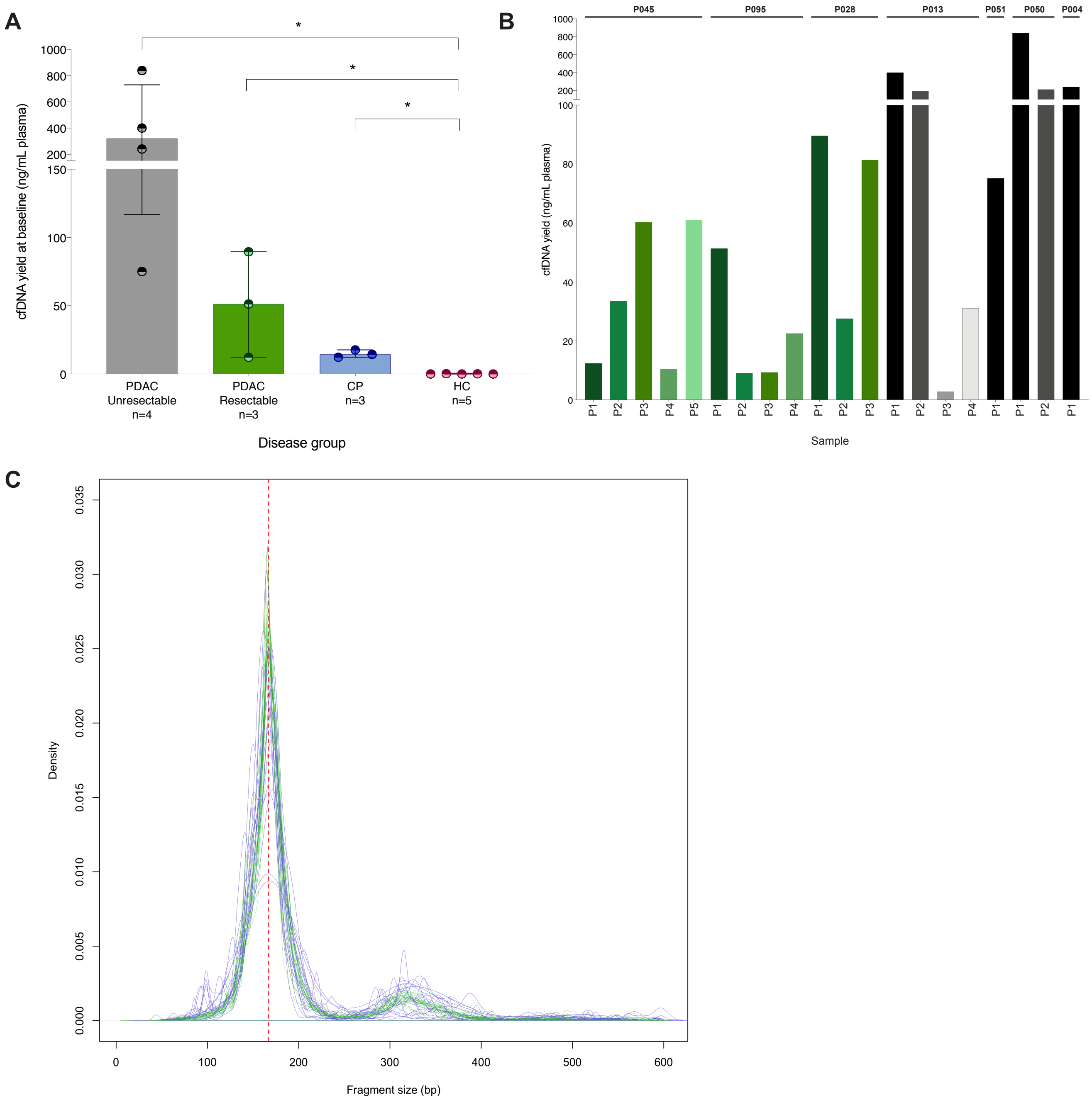


**Supplementary figure 1.** Isolated yields of cfDNA at baseline (pre-treatment) sampling in PDAC and control groups are shown in **(A)**. Mann-Whitney U tests were performed for comparison (*P $\leq$ 0.05). Yields of overall cfDNA in PDAC cases ranged from 12.34ng/mL to 840ng/mL plasma at P1 sampling. Extracted cfDNA yields from baseline (P1) and subsequent follow-up samples (P2-P5) from sequenced PDAC cases are shown in **(B)**. Fragmentation profiles of plasma sequencing reads from all n=20 samples in our cohort containing mutant *(purple)* and wild-type *(green)* alleles at target loci for candidate tumour mutations, as identified using our pipeline, are shown in **(C)**. A vertical *red* line indicating the modal 167bp mononucleosomal fragment size is shown on the graph.

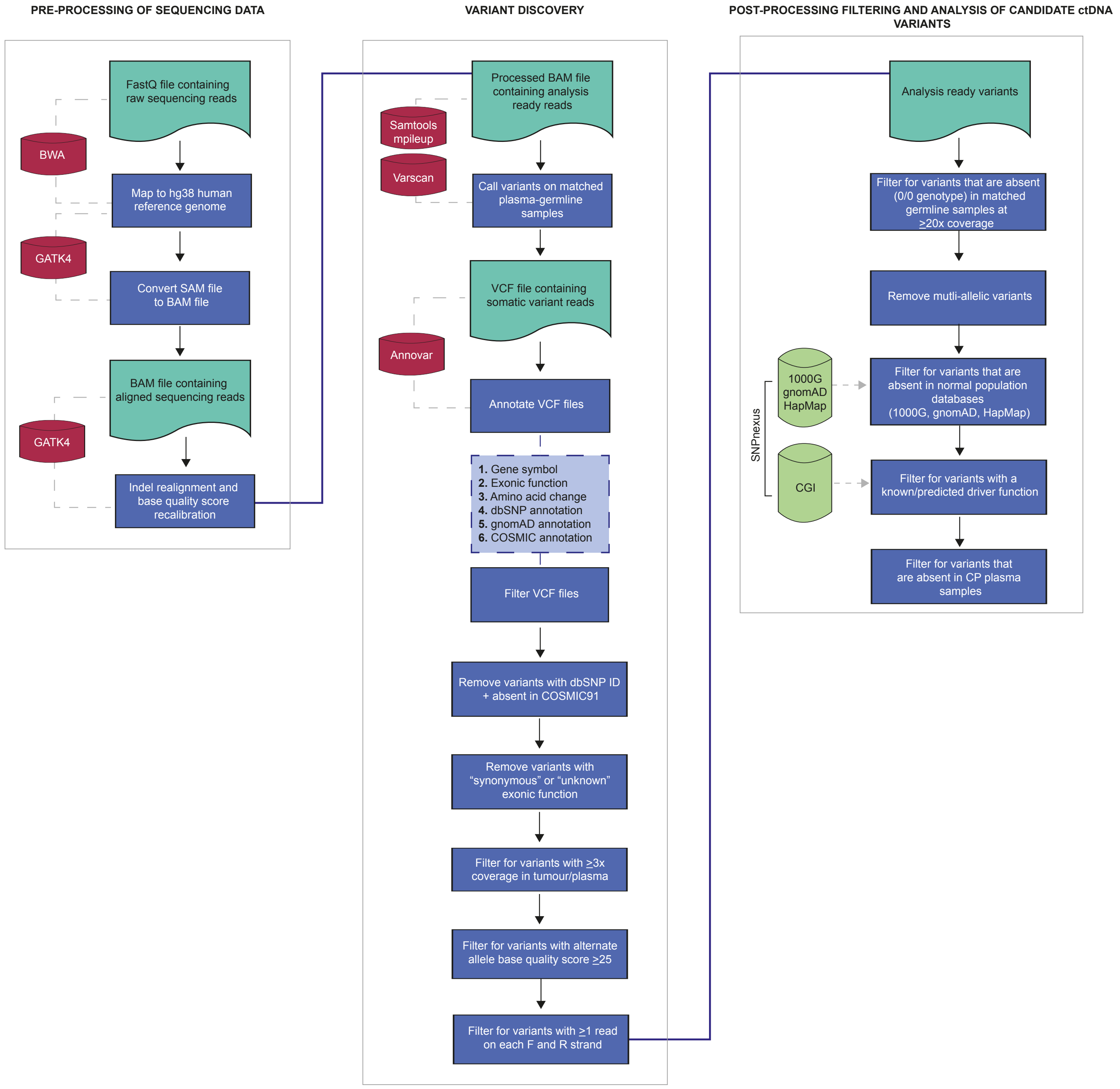


**Supplementary figure 2.** Summary of analytical pipeline used for the processing and analysis of plasma sequencing reads for identification of candidate ctDNA variants.

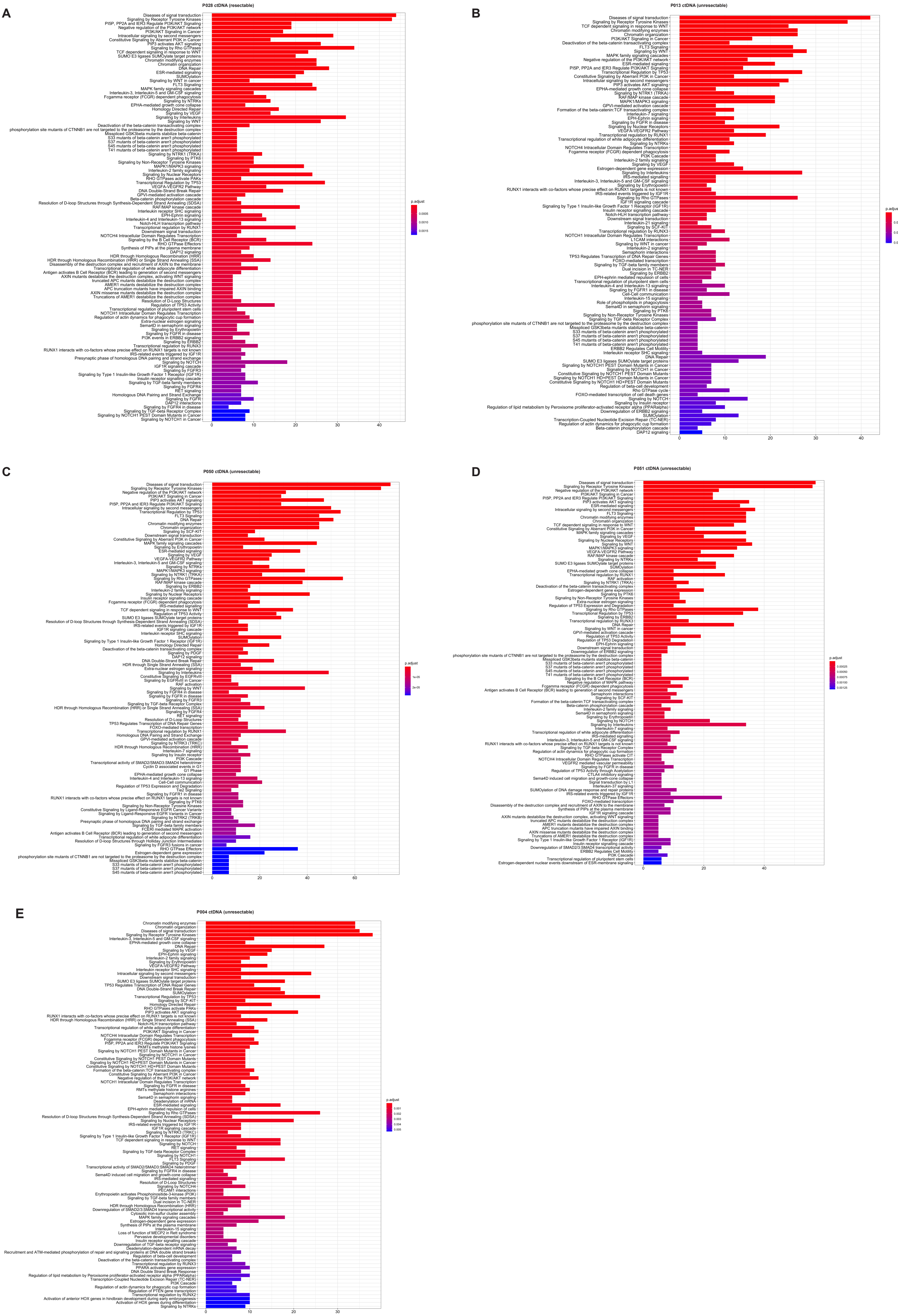
A
P028 ctDNA (resectable)
B
P013 ctDNA (unresectable)
C
P050 ctDNA (unresectable)
D
P051 ctDNA (unresectable)
E
P004 ctDNA (unresectable)
p.adjust

**Supplementary figure 3.** Enriched gene signalling pathways *(Reactome)* amongst ctDNA variants from patients 28 **(A),** 13 **(B),** 50 **(C),** 51 **(D)** and 04 **(E).** Multiple aberrations were observed in ctDNA within signalling pathways representative of PDAC, with frequent mutations in genes associated with TGF-$\beta$, WNT, NOTCH signalling and chromatin modification.

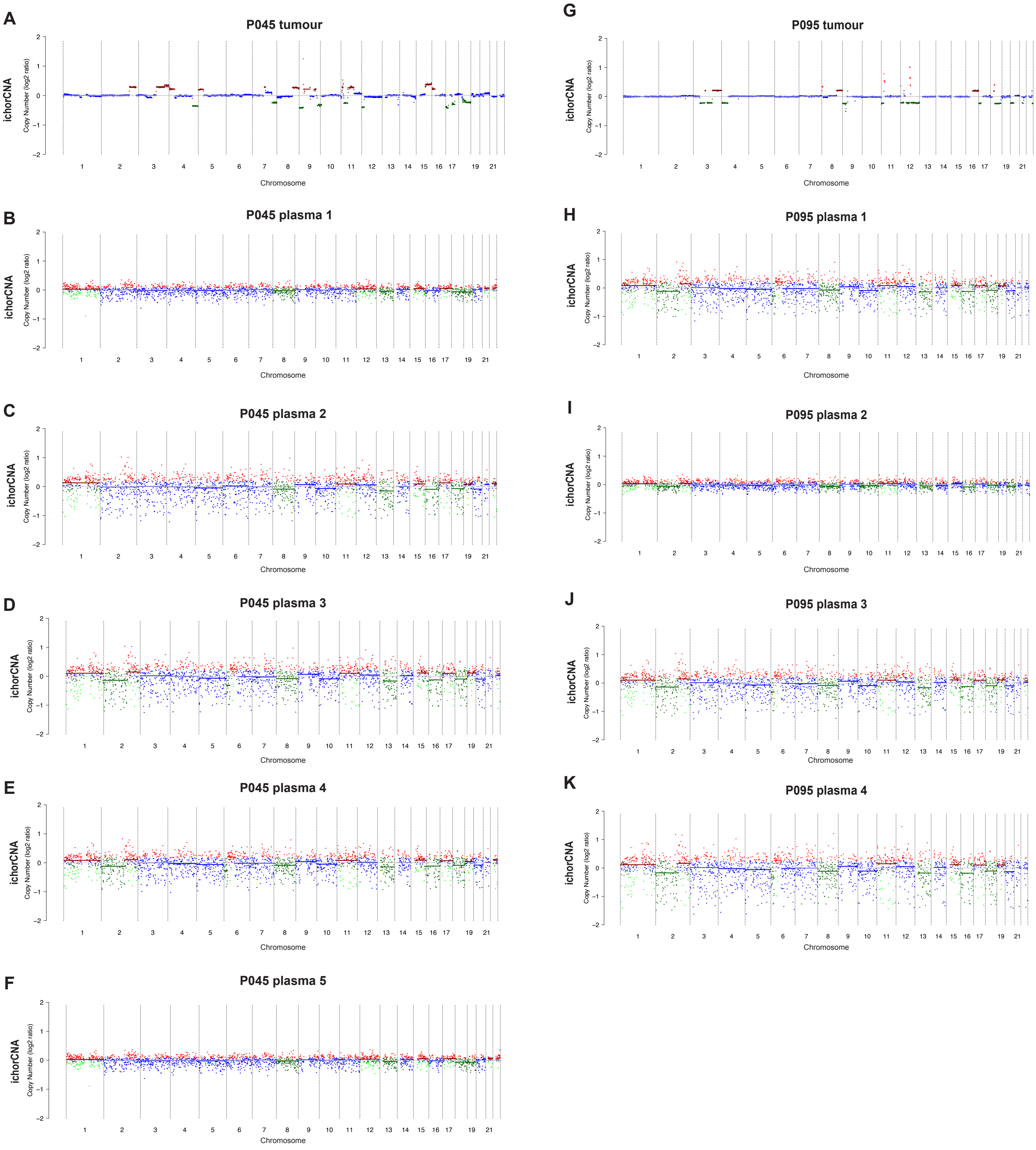


**Supplementary figure 4.** Genome-wide somatic copy number profiles (as derived from ichorCNA) in tumour and plasma samples from patients 45 **(A-F)** and 95 **(G-K)**.

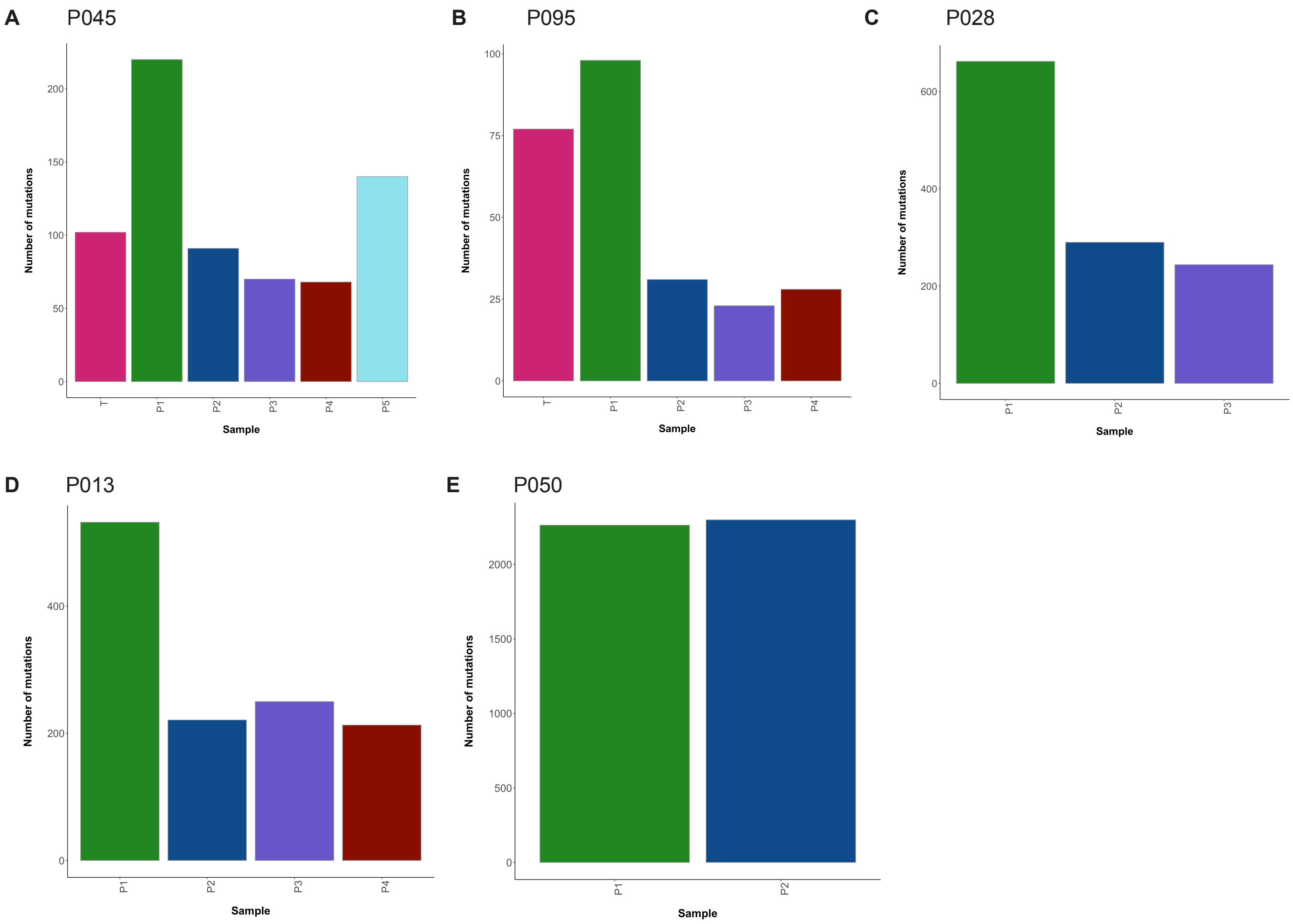


**Supplementary figure 5.** Bar plots showing the overall number of ctDNA mutations, (with known/predicted driver classifications) identified throughout serial plasma timepoints in patients with ≥2 plasma samples **(A-E)**. The number of ctDNA mutations varied significantly across sampled timepoints from individual patients. In all resectable patients **(A-C)**, a reduction in the total number of ctDNA mutations was observed following surgical removal of primary tumour lesions (P1 to P2 sampling). Similarly, reductions in the number of ctDNA mutations were observed in unresectable patient 13, during the course of first-line chemotherapy treatment (P1 to P2).

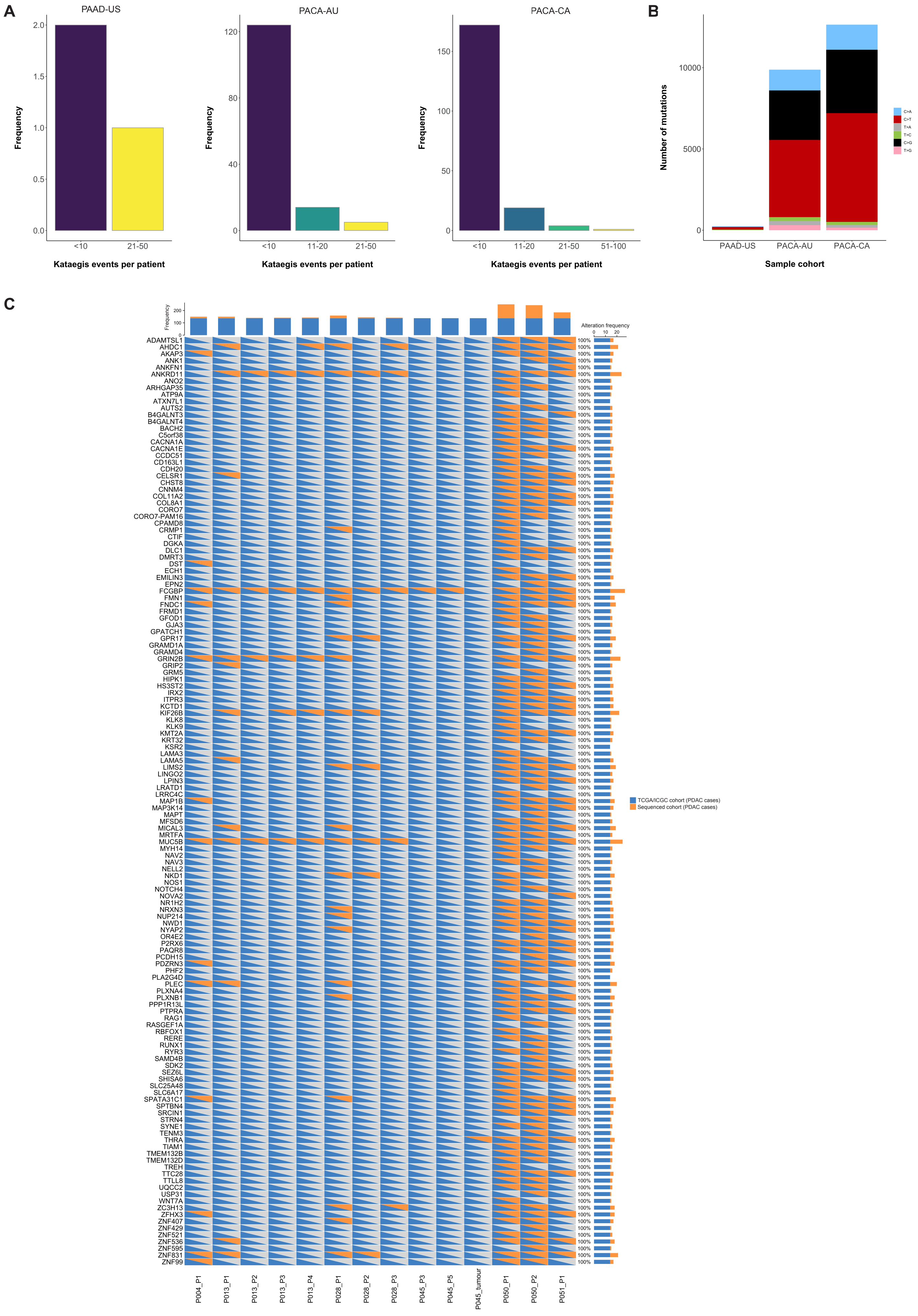
A
PAAD-US
PACA-AU
PACA-CA
Frequency
Kataegis events per patient
<10
11-20
21-50
51-100
B
Number of mutations
Sample cohort
PAAD-US
PACA-AU
PACA-CA
C>A
C>T
T>A
T>C
C>G
T>G
C
Alteration frequency
TCGA/ICGC cohort (PDAC cases)
Sequenced cohort (PDAC cases)
P004_P1
P013_P1
P013_P2
P013_P3
P013_P4
P028_P1
P028_P2
P028_P3
P045_P3
P045_P5
P045_tumour
P050_P1
P050_P2
P051_P1

**Supplementary figure 6.** Analysis of kataegis in TCGA/ICGC PDAC tumours. **(A)** Bar plots showing the total number of kataegis events detected in available tumour sequencing data from each cohort, using MAFtools. **(B)** Base substitution profiles of somatic mutations detected within regions of kataegis in each cohort are displayed. **(C)** Oncoprint displaying 137 genes that were found to harbour kataegis events across PDAC samples sequenced as part of the current study cohort *(orange)* and PDAC tumours from TCGA/ICGC cohorts *(blue)*. Kataegis events co-localising with *ERBB2* were not detected in TCGA/ICGC PDAC tumours.

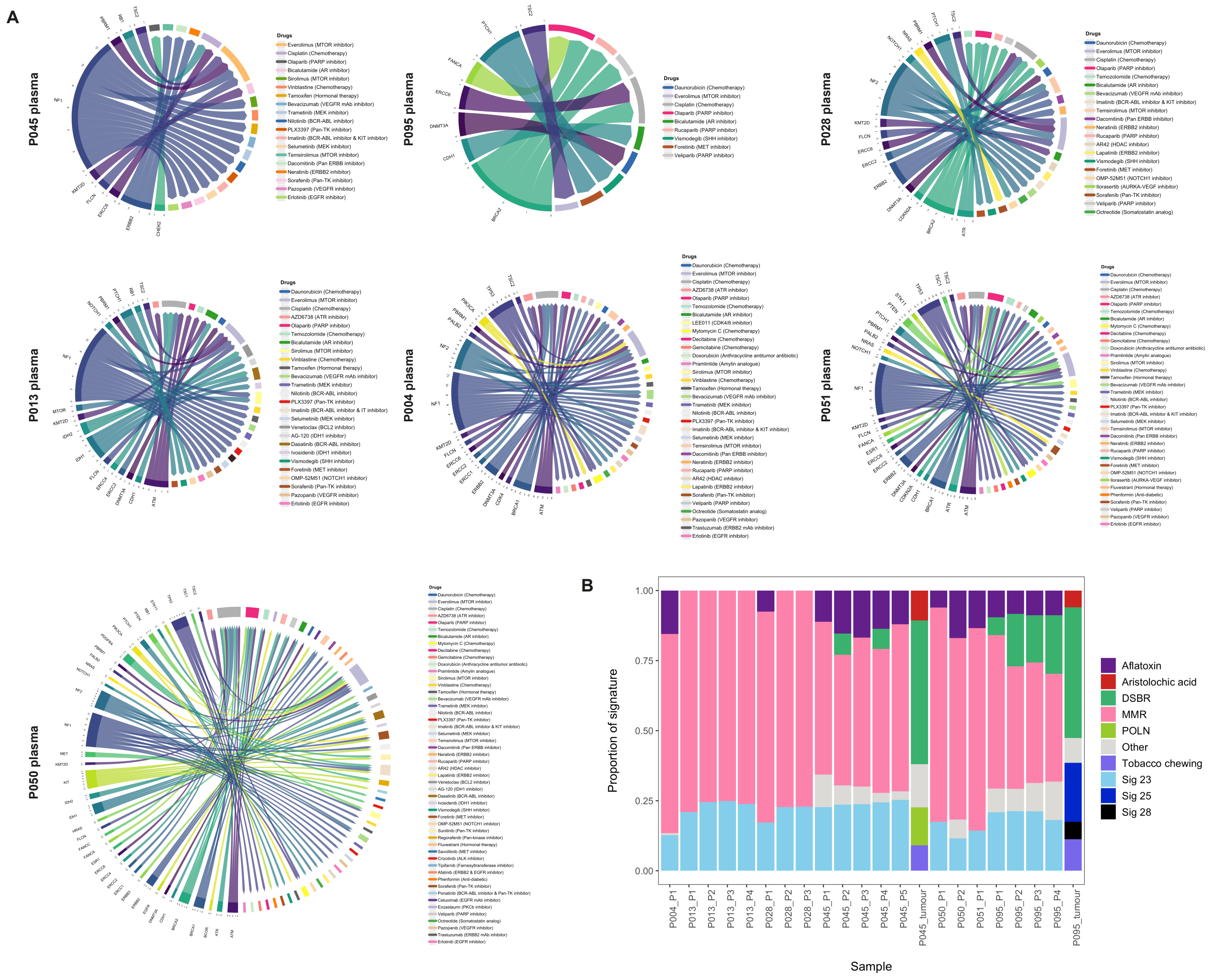


**Supplementary figure 7.** Examples of ctDNA genes containing driver mutations that were predicted to confer response to existing clinical/pre-clinical treatments using *in silico* predictive algorithms from *Cancer Genome Interpreter,* are shown. The *widths* of gene segments correspond to the number of unique drug targets identified for ctDNA alterations detected within that gene. **(B)** Bar plot displaying enriched (COSMIC) mutational signatures across sequenced tumour and plasma samples. The contribution of each signature as a proportion of total signatures detected in each sample is shown. Overall, 9 COSMIC signature classes were resolved in this cohort, including 3 signatures with currently unknown aetiologies *(Signature 23, Signature 25, Signature 28)*.

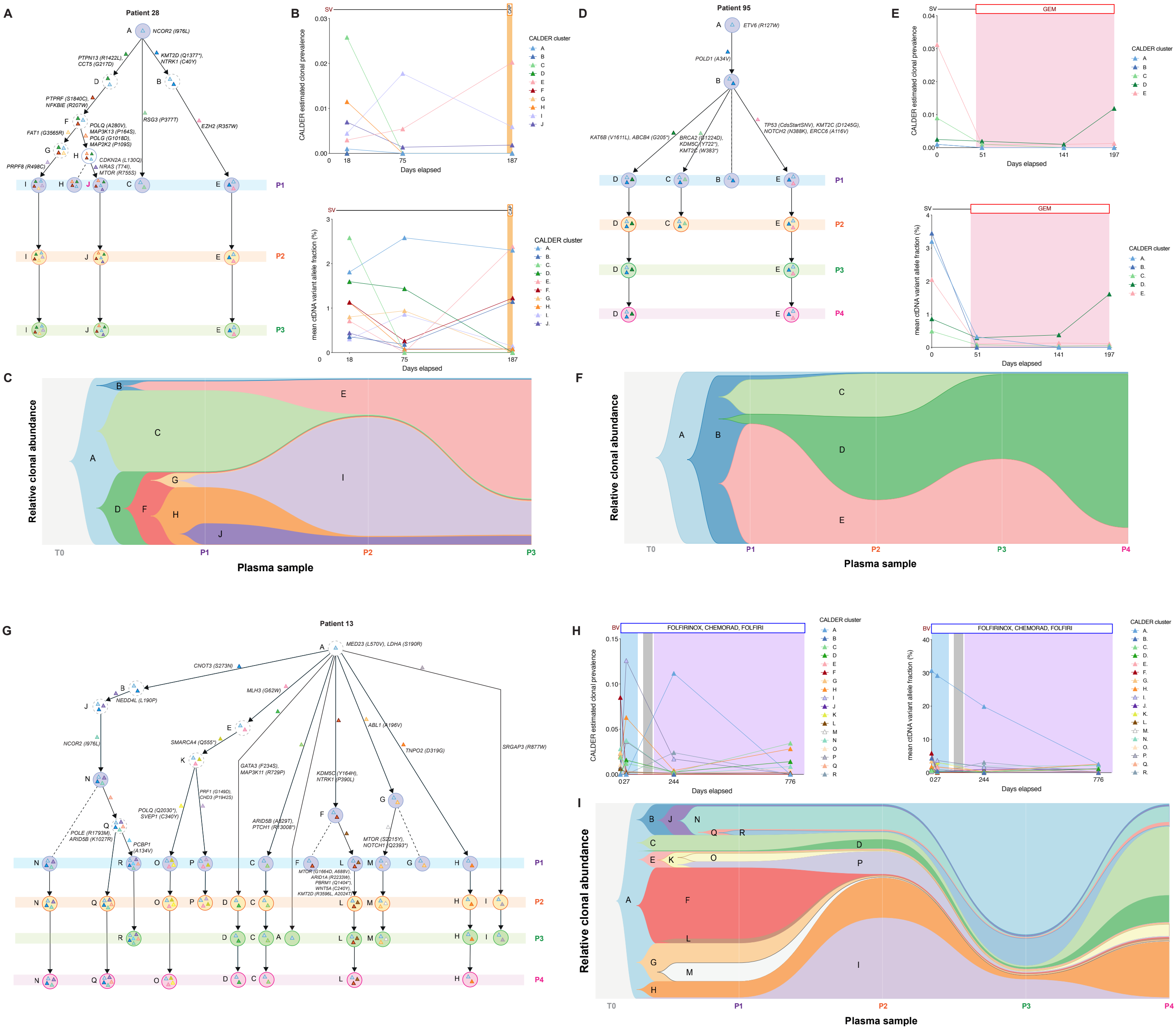

**Supplementary figure 8.** Analysis of clonal dynamics and evolutionary trajectories in patients 28 **(A-C)**, 95 **(D-F)** and 13 **(G-I)**. Longitudinally observed phylogenetic trees showing the predicted clonal evolutionary trajectories of individual ctDNA clones from each patient are shown in **(A)**, **(D)** and **(G)**. Scatterplots showing the total estimated prevalence of inferred clones in ctDNA across sampled timepoints, are shown alongside plots displaying the mean variant allele fractions of ctDNA mutations within each inferred clone in **(B)**, **(E)** and **(H)**. Clonal diagrams of the tree structures from (A), (D) and (G) are displayed in **(C)**, **(F)** and **(I)**.
