## Supplementary Table 1 for "Longitudinal profiling of circulating tumour DNA for tracking tumour dynamics in pancreatic cancer"

**Supplementary Table 1. Summary of the clinical characteristics of the study cohort**

| Patient ID | Sex | Ethnicity | Tumour location | Tumour grade | TNM at diagnosis | Site of metastases at diagnosis | Survival status | Recurrent/progressive disease |
|---|---|---|---|---|---|---|---|---|
| **P045** | Female | Caucasian: Western European | Head of pancreas | Poorly differentiated | pT3N0M0 | None | Alive | No recorded recurrence |
| **P095** | Female | Caucasian: Western European | Head of pancreas | Moderately differentiated | pT1N1M0 | None | Deceased | Hepatic metastases detected 6 months after baseline visit |
| **P028** | Male | Caucasian: Western European | Head of pancreas | Moderately differentiated | pT2N1M0 | None | Alive | No recorded recurrence |
| **P050** | Male | Caucasian: Western European | Head of pancreas | Moderately differentiated | T4N1M0 | None | Deceased | Increase in primary tumour volume observed 2 months after baseline visit |
| **P013** | Male | Caucasian: Western European | Head of pancreas | Poorly differentiated | T4N0M0 | None | Alive | No recorded progression |
| **P051** | Male | Afro-Caribbean | Head of pancreas | Poorly differentiated | T4N0M0 | None | Deceased | Hepatic metastasis 14 months after diagnosis |
| **P004** | Male | Afro-Caribbean | Pancreatic tail | Poorly differentiated | T3N1M1 | Liver | Deceased | Progression soon after diagnosis- |
